# Quantification of the IgG antibody response half-life for hybrid immunity to SARS-CoV-2

**DOI:** 10.1101/2025.07.29.25332411

**Authors:** Felicia Bongiovanni, Eamon Conway, Lauren Smith, Ramin Mazhari, Nicholas Kiernan-Walker, Emily Eriksson, Jodie McVernon, Ivo Mueller

## Abstract

**Background:** A firm understanding of SARS-CoV-2 hybrid immunity is crucial for our ongoing efforts to protect people from severe and fatal disease and assess population vulnerability to emerging novel variants. As many components of the immune response are unobserved and complex to investigate, some ambiguities and unanswered questions remain about hybrid immunity to COVID-19, such as the duration of the antibody response in individuals.

**Methods:** To address this, we evaluated longitudinal data that spanned up to 21 months from 53 SARS-CoV-2 naive individuals and 89 SARS-CoV-2 recovered individuals. We further separated individuals according to whether they received an mRNA or non-mRNA vaccine for their primary two-dose vaccinations. A hierarchical Bayesian framework was used to fit the parameters of single-phase exponential decay and bi-phasic exponential decay models to the observed data to estimate the magnitude and half-life of the spike-specific and receptor-binding domain (RBD)-specific IgG responses.

**Results:** Results from both our single-phase and bi-phasic exponential decay models estimate that the median half-life of the spike-specific and RBD-specific IgG response in individuals with hybrid immunity is almost double that of naive individuals who were only vaccinated. Recovered individuals who received an mRNA vaccine for their primary two-dose schedule were estimated to have the longest IgG half-life response in both model results. Through the single-phase exponential decay estimates, recovered mRNA recipients had a median IgG response half-life of 499 days (95% CrI: [404.9, 635.8]) to RBD antigen and 452.6 days (95% CrI: [376.4, 556.3]) to spike antigen. Estimates from the bi-phasic exponential decay model show that recovered individuals who received mRNA vaccinations had a median IgG response half-life of 859.1 days (95% CrI: [685.7, 1095.4]) to spike antigen and 807.8 days (95% CrI: [645.4, 1030.9]) to RBD antigen. Our results show that, across different model assumptions, individuals with hybrid immunity have an IgG response half-life that is considerably greater than that of individuals with only infection- or vaccine-induced immunity.

**Conclusions:** Our work provides important insight into the longevity of the IgG response to SARS-CoV-2 in individuals with hybrid immunity and can guide effective immunisation approaches to maintain and improve population-level protection.

## 1. Background

Since the emergence of SARS-CoV-2 in December 2019, extensive research has been conducted worldwide to understand the immune response to the virus. Despite this, our knowledge of the development and persistence of immunity to COVID-19 remains incomplete. Vaccines are primarily used to maintain or enhance immunity against pathogens, including SARS-CoV-2. However, vaccination is not always known to produce immune responses comparable to those elicited by natural infections [1–4]. In particular, numerous studies report that vaccination-acquired immunity is less durable and protective compared to infection-associated immunity [1, 5–8]. However, other studies suggest that there are no significant differences in protection against severe disease and the risk of reinfection in fully vaccinated individuals compared to those who have previously been infected [9–13]. Furthermore, additional studies demonstrate that hybrid immunity (achieved through a combination of natural infection and vaccination) provides more robust immunity against other variants of SARS-CoV-2 and more durable immunity than natural infection or vaccination alone [7, 12, 14–16]. These contrasting findings underscore that much remains to be understood about immunity to SARS-CoV-2 acquired by vaccination and natural infection.

The magnitude of the antibody response to SARS-CoV-2 depends on factors such as the type of vaccine received [17, 18], whether the exposure is due to vaccination or natural infection, and an individual’s exposure history. A significant gap in our knowledge is the duration of protection against SARS-CoV-2 mediated by antigen-specific antibodies [19]. As antibodies, especially the IgG isotype, are well known to be correlated with protection against disease [20–23], a significant area of interest is to determine the duration of the antibody response. Long-term humoral immunity against SARS-CoV-2 is maintained by long-lived plasma cells (LLPCs) that continuously secrete antibodies for months to years and memory B cells that maintain immunological memory to mount rapid and robust responses upon re-exposure. Cells such as LLPCs that are critical to protective immunity remain understudied due to the challenges in investigating this cell population without invasive procedures given their residence in the bone marrow [24]. Additionally, given that hybrid immunity has become the dominant immune status, it is increasingly complex to ascertain an individual’s exposure history. Thus, a comprehensive assessment of the duration and strength of the antibody response elicited by natural exposure and vaccination to SARS-CoV-2 is challenging to determine using data from the current population.

In contrast to LLPCs, antibody titres are readily available in the blood and can be measured to provide insight into the underlying dynamics of humoral kinetics through mathematical modelling [25–27]. Many previous studies modelling the antibody response to SARS-CoV-2 antigens explain kinetics through one-phase exponential decay models and linear regression [28–34]. These are limiting to our understanding of immunity to COVID-19 as they overlook essential components of antibody maintenance and do not account for differences in short- and long-lived immune dynamics. Other studies account for two-phase antibody decay through power-law models [35], gamma models [36], and bi-phasic exponential decay models [37–40], accounting for the contribution of SLPCs and LLPCs to antibody dynamics. Of these studies, none report on the interface between mRNA and non-mRNA vaccine-acquired immunity, naturally acquired immunity, and how this translates to hybrid immunity. This information is crucial to inform long-term recommendations on global vaccine uptake, scheduling vaccine administration, and better protecting and preparing the population against future pandemics caused by novel viruses. In this study, we aimed to quantify the magnitude and duration of the IgG antibody response to the SARS-CoV-2 spike and receptor-binding domain antigens using longitudinal serological data from Victoria, Australia and an antibody kinetics model that accounts for bi-phasic exponential decay. The data from Australia presented a unique resource for investigating immunity to COVID-19 in a controlled, real-world setting, given the strict lockdown procedures implemented and the limitations on international travel within the country. These factors created a low-transmission early population cohort that can be studied without a significant risk of frequent or undetected SARS-CoV-2 infections influencing the results. Additionally, we compare the bi-phasic exponential decay model to a single exponential decay model to highlight the importance of model assumptions and their impact on evaluating immunity. We considered the differences between mRNA and non-mRNA vaccine types, as well as the infection history of individuals, to report on the effect of vaccine-acquired immunity and naturally acquired immunity in achieving hybrid immunity.

## 2. Materials and Methods

### 2.1 Study population

The SARS-CoV-2 study population comprises data collected from individuals recruited as part of the COVID PROFILE study [41], which was launched in Melbourne, Victoria, in October 2020. The recruitment period for the study was from October 7, 2020, to July 5, 2022. Most participants were enrolled between October 2020 and June 2021, and the study sample collection and follow-up concluded in May 2023. Adult community members were recruited through advertising campaigns in primary care networks, tertiary teaching hospitals, and online social media platforms. A total of 178 individuals were enrolled in the study, of which seven participants were excluded from the study as they failed to provide a baseline sample, as required by the study schedule. The study inclusion criteria considered only non-pregnant individuals over 18 years of age.

Individuals who were SARS-CoV-2 naive (*n* = 73) and those who had recovered from SARS-CoV-2 infection (*n* = 98) were enrolled, with all participants providing written informed consent as per the Declaration of Helsinki. The study was approved by the Walter and Eliza Hall Institute (Projects 20/08) and the Melbourne Health Human Research Committees (RMH69108). Most participants received at least two doses of a SARS-CoV-2 vaccine. The majority of individuals in both the SARS-CoV-2-naive and SARS-CoV-2-recovered groups received either the Pfizer–BioNTech mRNA vaccine (BNT162b2) or the AstraZeneca adenoviral vector vaccine (ChAdOx1-S), with a minor subset of participants who received either the Moderna mRNA vaccine (mRNA-1273) or the Novavax virus-like particle vaccine (NVX-CoV2373). All vaccines contained the spike antigen of the ancestral variant. A subset of vaccinated individuals also received up to five doses, most of which were booster vaccinations with BNT162b2 or CX-024414. Clinical and experimental data were available from each participant, including their date of diagnosis and vaccination, symptoms, and longitudinal antibody data.

#### 2.1.1 Serology data

Antibody levels were measured in plasma collected from individuals at their enrolment date and, in most cases, with multiple follow up sample collections until either the conclusion of the study or at the individual end date. Samples were collected at enrollment, two weeks after any vaccination or natural infection, and then every three months after vaccination or infection. There was a median of 9 samples collected for naive individuals and a median of 11 samples collected for recovered individuals (Figure S1). IgG antibodies that recognise and bind to the full-length spike and receptor-binding domain (RBD) of SARS-CoV-2 were measured using a Luminex MAGPIX multiplex assay [42]. All antigens were based on the ancestral variant sequence. Antibody levels for each participant were reported in mean fluorescence intensity (MFI) and adjusted for plate-to-plate variation using a five-parameter logistic regression model algorithm based on standard curves and quality control samples.

### 2.2 Antibody kinetics model

#### 2.2.1 Single exponential decay model

We employed an exponential decay model to describe the waning of antibodies (*A*_*S*_) over time *t*, assuming that decay occurs at a single constant rate,*λ*. This is described by,

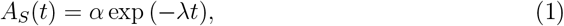

where *σ* is the initial antibody level at *t* = 0. The decay rate *λ* is related to the half-life of the antibody response, calculated through

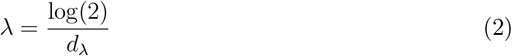

where *d*_*λ*_ is the half-life of the antibody response measured in days.

#### 2.2.2 Bi-phasic exponential decay model

A previously published mathematical model describing the generation and bi-phasic waning of antibodies [25, 43, 44] was used to capture the kinetics of short-lived plasma cells (*S*), long-lived plasma cells (*L*), and antibodies (*A*_*B*_) over time *t*. These are governed by a system of ordinary differential equations (ODEs),

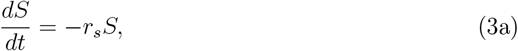

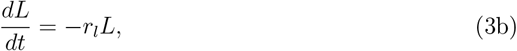

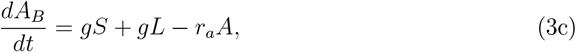

where *r*_*s*_, *r*_*l*_, and *r*_*a*_ are the decay rates over days^−1^ of short-lived plasma cells, longlived plasma cells, and antibodies, respectively and *g* is the antibody secretion rate of both short-lived and long-lived plasma cells. The decay rates are related to the half-life of each immune component through

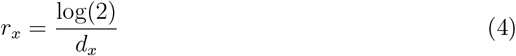

where *d*_*x*_ is the half-life of the *x*th immune component and *x* ∈ {*s, l, a*}.

The initial conditions that accompany Equation 3 are defined as

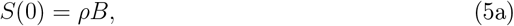

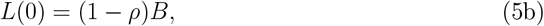

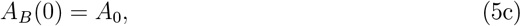

in which *B* is the total increase of plasma cells, *ρ* is the proportion of the increase that is short-lived plasma cells, and *A*_0_ is the pre-existing level of antigen-specific antibodies.

This system of ODEs can be solved to give the total antibody titres through time,

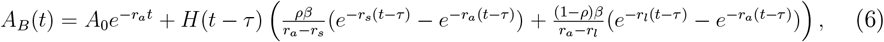

to capture the production and bi-phasic decay of antibodies at time *t* following exposure to an antigen at time *τ*. As *g* and *B* are non-identifiable parameters we estimate *β*= *gB*. The Heaviside function denoted by *H* specifies that a boost of antibodies only occurs if *t* ≥ *τ*. It is important to note that we assume that any pre-existing antibodies that are still present are being maintained by LLPCs. Therefore, we assume that they decay exponentially at rate *r*_*l*_, changing the expression in Equation 6 to

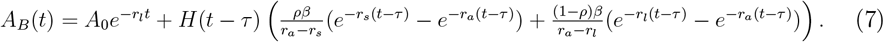

As an individual can have multiple exposures over a follow-up period, we account for multiple *τ* values. We also want to evaluate the magnitude of the antibody response across exposures, and introduce a parameter *m* to measure the fold change in *β* over the follow-up period for an individual. This leaves us with the final equation,

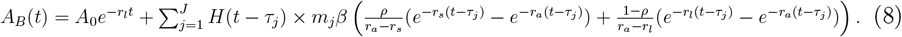

### 2.3 Fitting model parameters to data

#### 2.3.1 Single exponential decay model

We used a Bayesian hierarchical modelling approach to estimate *d*_*λ*_ for each individual 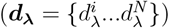 and at the population level 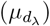. We define the expression to estimate these parameters from observed antibody measurements ***y*** over known time points for *N* individuals as,

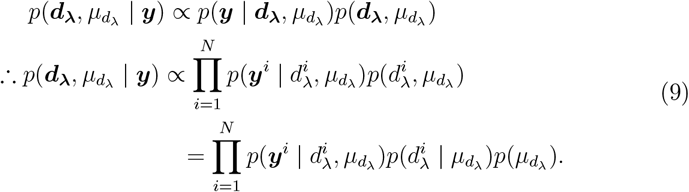

We describe the measurement of ***y***^*i*^ for the *i* -th individual to follow,

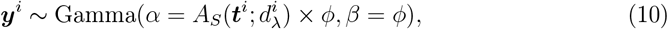

where ***t***^*i*^ are the observed time points. Gamma denotes the Gamma distribution classically parameterised by shape *ε* and rate*β*. A Gamma distribution was deemed suitable given that the error variance of the MAGPIX instrument, used to measure the antibody titres, is proportional to the magnitude of its reading. As we want the shape of the distribution to be centered around the expected value of the model-predicted antibody level, we determine *σ* by calculating,

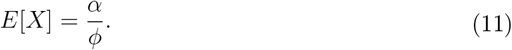

As we have *N* individuals, each with a varying number of time points annotated by *T*, we state the likelihood to be,

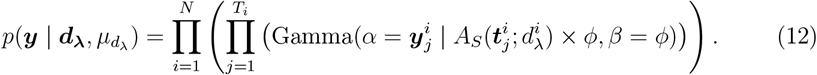

We choose an appropriate prior for *ϕ* to be the standard lognormal distribution,

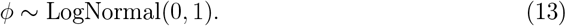

Given that *d*_*λ*_ is expected to be positive, we selected a lognormal distribution as an appropriate prior distribution. We want the mean of the distribution to be centered around the expected value for each prior, so we solve for *μ* given,

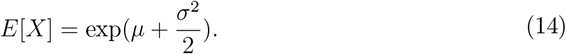

We define a function for the solved expression in Equation 14 as,

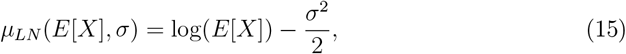

to simplify the notation of the calculation of the mean of the lognormal. Thus, our prior for the individual-level parameters is expressed by,

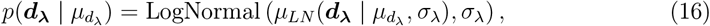

where *σ*_*λ*_ also has a standard lognormal distribution for a prior. Lastly, we define our prior for 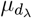 to be

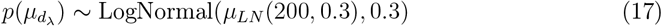

given the results of previous studies [28, 29, 31, 33].

By combining Equation 12 and Equation 16 into Equation 9, we get our final distri-bution of

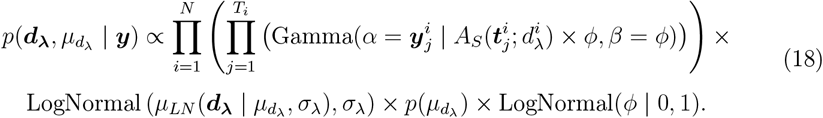

#### 2.3.2 Bi-phasic exponential decay model

We developed a Bayesian hierarchical modelling approach to estimate the model parameters from observed antibody measurements over known time points for *N* individuals. To do so, we defined the expression,

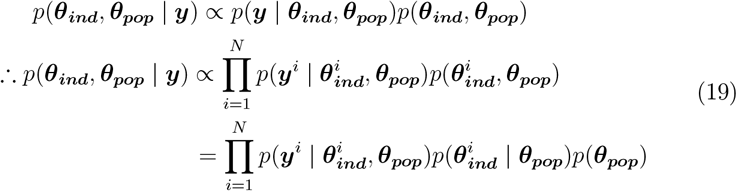

where 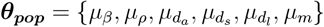 is the vector containing the population-level model parameters and 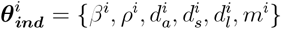 contains the individual-level model parameters for individual *i*. The parameters *d*_*a*_, *d*_*s*_, and *d*_*l*_ are the half-lives for antibodies, shortlived plasma cells, and long-lived plasma cells, respectively. The vector ***y*** represents the observed antibody levels over time for the entire cohort.

We describe the measurement of ***y***^*i*^ for the *i* -th individual to follow,

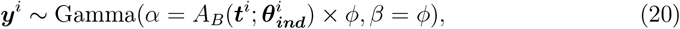

where ***t***^*i*^ are the observed time points. Gamma denotes the Gamma distribution classically parameterised by shape *α* and rate *β*. To avoid confusion with the model parameter *β* in Equation 8, the Gamma rate parameter will be referred to as*ϕ*. As with the other model, we want the shape of the distribution to be centered around the expected value of the model-predicted antibody level, following Equation 11.

As we have *N* individuals, each with a varying number of time points annotated by *T*, we state the likelihood to be,

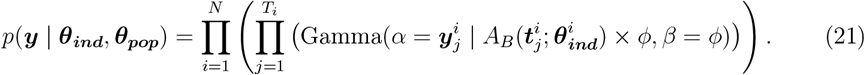

We once again choose an appropriate prior for *ϕ* to be the standard lognormal distribution,

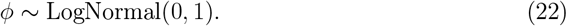

Furthermore, we define the individual-level priors for ***θ***_*ind*_ as,

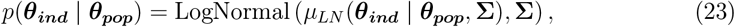

where = diag(***X***) and 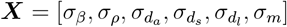. We consider a standard lognormal distribution to be an appropriate prior for ***X***, giving,

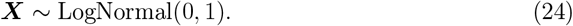

We assume that there are no pre-existing antibodies for SARS-CoV-2 for all participants in the COVID PROFILE study prior to their first recorded exposure (first vaccination for the naive cohort, first natural infection for the recovered cohort); therefore, we assume *A*_0_ = 1 (due to the bounds of the Gamma distribution, it cannot be set to 0). The specific priors chosen for ***θ***_*pop*_ are described in Table 1 below.

**Table 1:** Population-level priors for the IgG response to spike and receptor-binding domain protein.

| Parameter | Description (unit) | Prior | Source |
| --- | --- | --- | --- |
| $A_0$ | The level of pre-existing antibodies (adjusted MFI) | 1 | Arbitrary |
| $\mu_\beta$ | The increase in antibodies post-exposure (adjusted MFI) | $\text{LogNormal}(\mu_{LN}(1500, 0.3), 0.3)$ | Arbitrary |
| $\mu_\rho$ | The proportion of antibody-secreting cells that are short-lived | $\text{Beta}(5, 2)$ | [37] |
| $\mu_{d_l}$ | The half-life of the long-lived plasma cells (days) | $\text{LogNormal}(\mu_{LN}(400, 0.2), 0.2)$ | [36, 37] |
| $\mu_{d_s}$ | The half-life of the short-lived plasma cells (days) | $\text{LogNormal}(\mu_{LN}(3.5, 0.15), 0.15)$ | [45–48] |
| $\mu_{d_a}$ | The half-life of the IgG antibodies (days) | $\text{LogNormal}(\mu_{LN}(21, 0.1), 0.1)$ | [49–52] |
| $\phi$ | The rate parameter of the Gamma distribution | $\text{LogNormal}(0, 1)$ | [53] |
| $\mu_m$ | The fold change in $\beta$ across exposures | $\text{LogNormal}(\mu_{LN}(5, 0.5), 0.5)$ | [54] |

By combining Equation 21 and Equation 23 into Equation 19, we get,

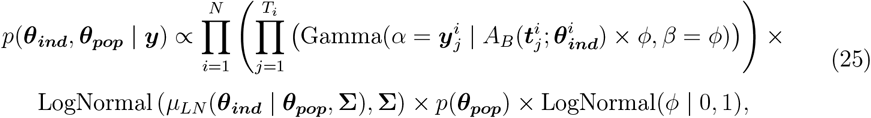

which is our final distribution.

#### 2.3.3 Inference

To sample from the posterior distribution we used Stan [55], a probabilistic programming language that supports the No-U-Turn sampler [56], an extension of Hamiltonian Monte Carlo [57]. Stan is used through RStan (version 2.21.8) [58], the R interface to Stan, within the R (version 4.3.1) environment [59].

Four chains with 10000 iterations each were run for the single exponential model, discarding 4000 iterations total for warm-up. This provided 36000 posterior samples for estimated parameters. All sampler parameters were left as the Stan default.

For the bi-phasic exponential model, four chains were run with 20000 iterations each, with 5000 discarded as warm-up. This produced 60000 posterior samples for each model parameter at the population-level and individual-level. Some of the Stan parameters that control the sampler were changed from the default to improve estimations. We set adapt_delta = 0.999 and max_treedepth = 12, while all other sampler parameters were left as the default values.

Traceplots and the effective sample size,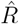, were evaluated to ensure that chains had converged and were well-mixed for both models.

### 2.4 Statistical Analysis

To test whether the difference in geometric means between the cohort and vaccination groups was significant, we conducted a t-test on the log-transformed adjusted MFI of the spike-specific and RBD-specific IgG titres. The mean of the log-transformed antibody levels was then converted to its exponential form to compare the values on their original scale. We calculated the posterior probability of the estimates of one model parameter being greater than another model parameter, or that the estimates between cohort or vaccine groups were greater, given

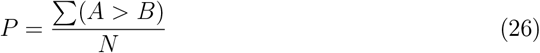

where *A* and *B* represent the estimates of group A and group B, respectively, and *N* is the total sample size.

## 3. Results

### 3.1 Summary of observed data

The COVID PROFILE data consisted of 171 individuals. Before conducting our modelling, we cleaned the data and removed 17 individuals with fewer than two antibody measurements. Additionally, we excluded any samples from individuals where an antibody measurement was not accompanied by a date of collection. We also removed four individuals who did not receive a vaccination and did not experience an infection, as they would have no information on the antibody response to exposure. Lastly, we removed four individuals who were vaccinated prior to enrolment in the study. These changes reduced the total sample size to 142 individuals, separated as 53 SARS-CoV-2-naive individuals and 89 SARS-CoV-2-recovered individuals. We separated both the naive and recovered cohorts based on the type of vaccine individuals received for their initial two doses. Those who received either BNT162b2 or mRNA-1273 were grouped into the mRNA vaccine group, and anyone who received ChAdOx1-S or NVX-CoV2373 was considered to be in the non-mRNA vaccine group. A summary of the groupings of individuals by cohort and initial vaccine received is presented in Table 2.

**Table 2:** Grouping of COVID PROFILE individuals.

| Initial vaccines | Naive cohort | Recovered cohort | Group |
| --- | --- | --- | --- |
| BNT162b2 (Pfizer-BioNTech) | 31 | 50 | mRNA |
| mRNA-1273 (Moderna) | 1 | 0 | mRNA |
| ChAdOx1-S (AstraZeneca) | 19 | 30 | non-mRNA |
| NVX-CoV2373 (Novavax) | 2 | 1 | non-mRNA |
| None | 0 | 8 | NA |

In addition, we identified individuals who experienced a natural infection during their participation in the study. Of those who naturally contracted SARS-CoV-2, all were infected after receiving their first two doses of vaccination. Individuals in the naive cohort experienced their infections between January 4, 2022, and January 15, 2023, coinciding with the Omicron wave (which occurred from the end of 2021 to the beginning of 2022). Those in the recovered cohort all experienced their first natural infection during the initial wave with the ancestral strain and experienced reinfections from October 7, 2021, to December 15, 2022. For any individuals naturally infected with SARS-CoV-2 during the study, we removed any observations after the exposure. The group with the highest number of individuals was the recovered cohort that received a full two-dose schedule of mRNA vaccinations, comprising a total of 50 people. Similarly, in the naive cohort, more people received mRNA vaccinations (*n* = 32) than non-mRNA vaccinations (*n* = 21). Table 3 also shows that there were more female participants in most groups than male participants and mRNA recipients were, on average, younger than non-mRNA recipients.

**Table 3:** Demographics of cohorts.

|  | Sex |  | Age | Cohort |
| --- | --- | --- | --- | --- |
|  | Male | Female | Median (Range) |  |
| **mRNA vaccine** |  |  |  |  |
| No infection | 11 | 21 | 36.5 (19 - 67) | Naive |
| Infected pre-vaccination | 14 | 36 | 40.5 (21 - 66) | Recovered |
| **non-mRNA vaccine** |  |  |  |  |
| No infection | 8 | 13 | 54 (25 - 84) | Naive |
| Infected pre-vaccination | 19 | 12 | 57 (23 - 75) | Recovered |
| **No vaccination** |  |  |  |  |
| Infected before study enrolment | 1 | 7 | 52 (21 - 63) | Recovered |

### 3.2 Comparing the observed IgG antibody response across cohorts

We observe the distribution and geometric mean of spike-specific and RBD-specific IgG antibodies produced in response to first vaccination, second vaccination, and booster vaccination in Figure 1. Anti-spike IgG measured 2-4 weeks after both the first and second vaccination in the recovered cohort had a higher geometric mean for both the mRNA and non-mRNA recipients than their respective counterparts in the naive cohort (p-value < 0.01, Table S1). We obtained similar results for anti-RBD IgG in previously infected non-mRNA recipients. However, we no longer observed a significant difference between the naive and recovered cohorts after individuals received a booster vaccination (p-value > 0.05, Table S1). This indicates that it took a complete two-dose primary vaccination schedule and a booster vaccination for individuals in the naive cohort to reach IgG antibody titres similar to those of individuals who had been previously infected. Individuals who received mRNA vaccinations in both the naive and recovered cohorts were observed to have higher IgG titres toward spike antigen after two vaccinations than non-mRNA recipients (p-value < 0.01, Table S2). We see a similar difference in response to RBD protein for previously infected mRNA recipients but not for naive recipients (p-value < 0.05, Table S2). Therefore, the results in Figure 1 suggest that a higher level of IgG titres is achieved through two doses of mRNA vaccination (being predominantly BNT162b2) than non-mRNA vaccination (predominantly ChAdOx1-S).

**Figure 1:**
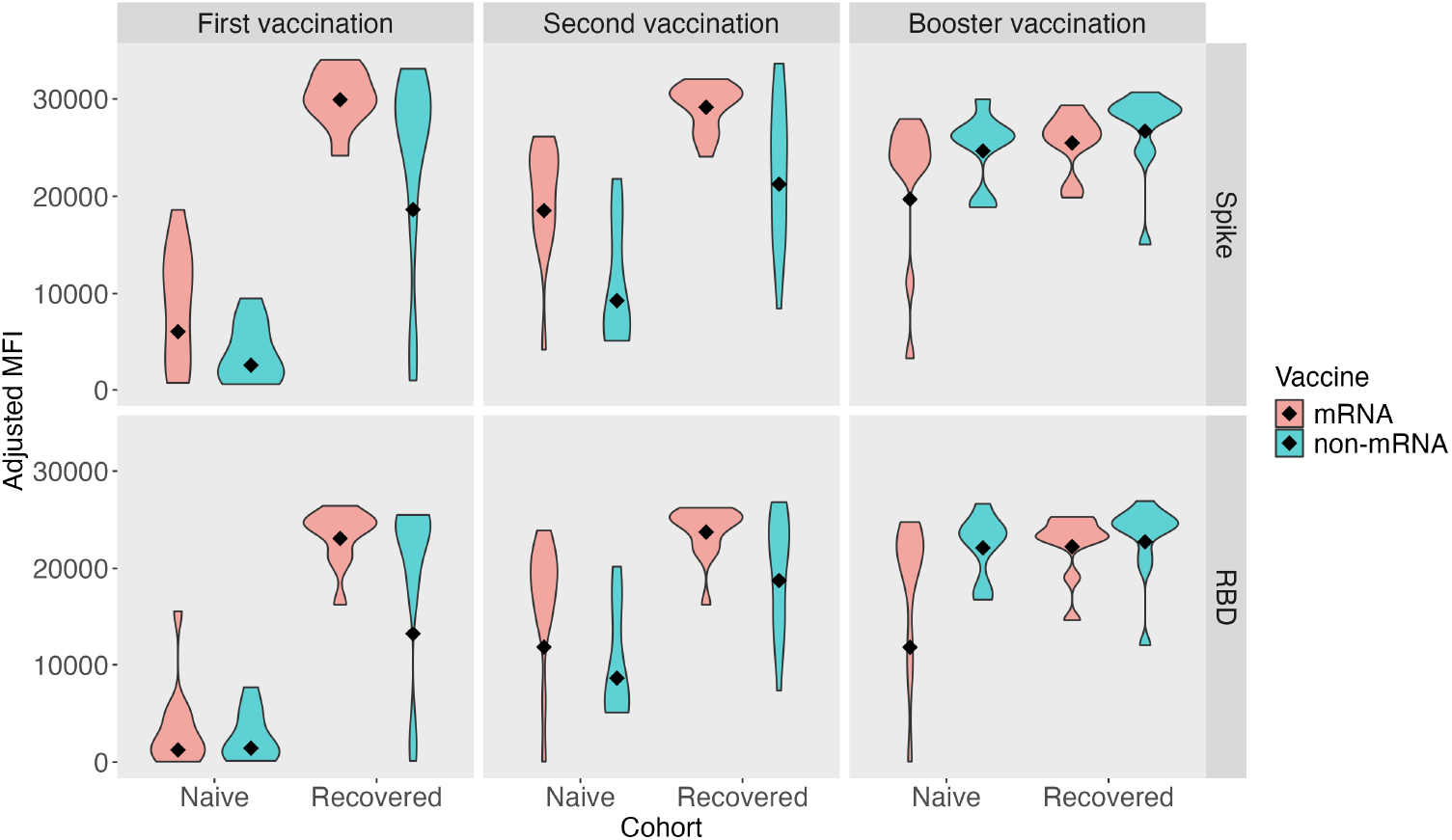
The IgG antibody levels to spike and RBD 2-4 weeks after receiving first vaccination, second vaccination, and booster vaccination across the naive and recovered cohorts. Violin plots illustrate the distribution of antibody titres measured in adjusted MFI and the diamonds signify the geometric mean for each group. Plots are separated by vaccination dose (top) and antigen response (right) and violin plots are separated according to cohort. The colours to represent the IgG levels from mRNA recipients (red) and non-mRNA recipients (blue) within each cohort.

In Figure 1, it is apparent that there is no substantial difference in the RBD-specific IgG levels of the recovered cohort as the number of vaccinations received increases (p-value > 0.05, Table S3). In response to the spike protein, this was only true between the first and second mRNA doses for recovered individuals, as there was a significant increase between the second vaccination and the booster dose (p-value < 0.05, Table S3). There was a significant increase in the spike-specific and RBD-specific IgG levels in naive individuals receiving the mRNA vaccination between their first and second dose, but not between their second vaccination and booster dose (p-value < 0.01, Table S3). In contrast, there was a significant increase in the IgG titres for naive non-mRNA recipients as the number of doses received increased (p-values < 0.01, Table S3). These results indicate that naive individuals reach their peak antibody titre after only two doses of an mRNA vaccine. However, a booster dose is required by naive individuals receiving non-mRNA vaccines to achieve an IgG level similar to that of mRNA recipients.

### 3.3 Analysing the increase in IgG antibodies upon exposure

Next, we fit parameters from both antibody kinetics models to the COVID PROFILE data to investigate the characteristics of the increase in IgG levels across exposures (interpreted by the bi-phasic decay model in Equation 8) and the decay of the IgG response (interpreted both by the bi-phasic decay model and the single decay model in Equation 1). The upper labels of Figure 2 describe the two parameters related to antibody production after exposure to the spike and RBD proteins, and the labels on the right specify the cohort and the type of vaccination received by individuals. We begin the analysis by examining the baseline of individuals from the recovered cohort who either did not receive a vaccination or by using antibody samples from individuals before they received any vaccination. The increase in anti-spike IgG antibodies per day post-exposure was estimated to be a median of 1383.02 adjusted MFI (95% CrI: [843.78, 2281.17]), and anti-RBD IgG antibodies had a median increase of 1370.72 adjusted MFI (95% CrI: [854.38, 2222.95]) after exposure to natural infection. As we only modelled the response to a single exposure, there is no estimate of the fold change in *β* (*μ*_*m*_).

**Figure 2:**
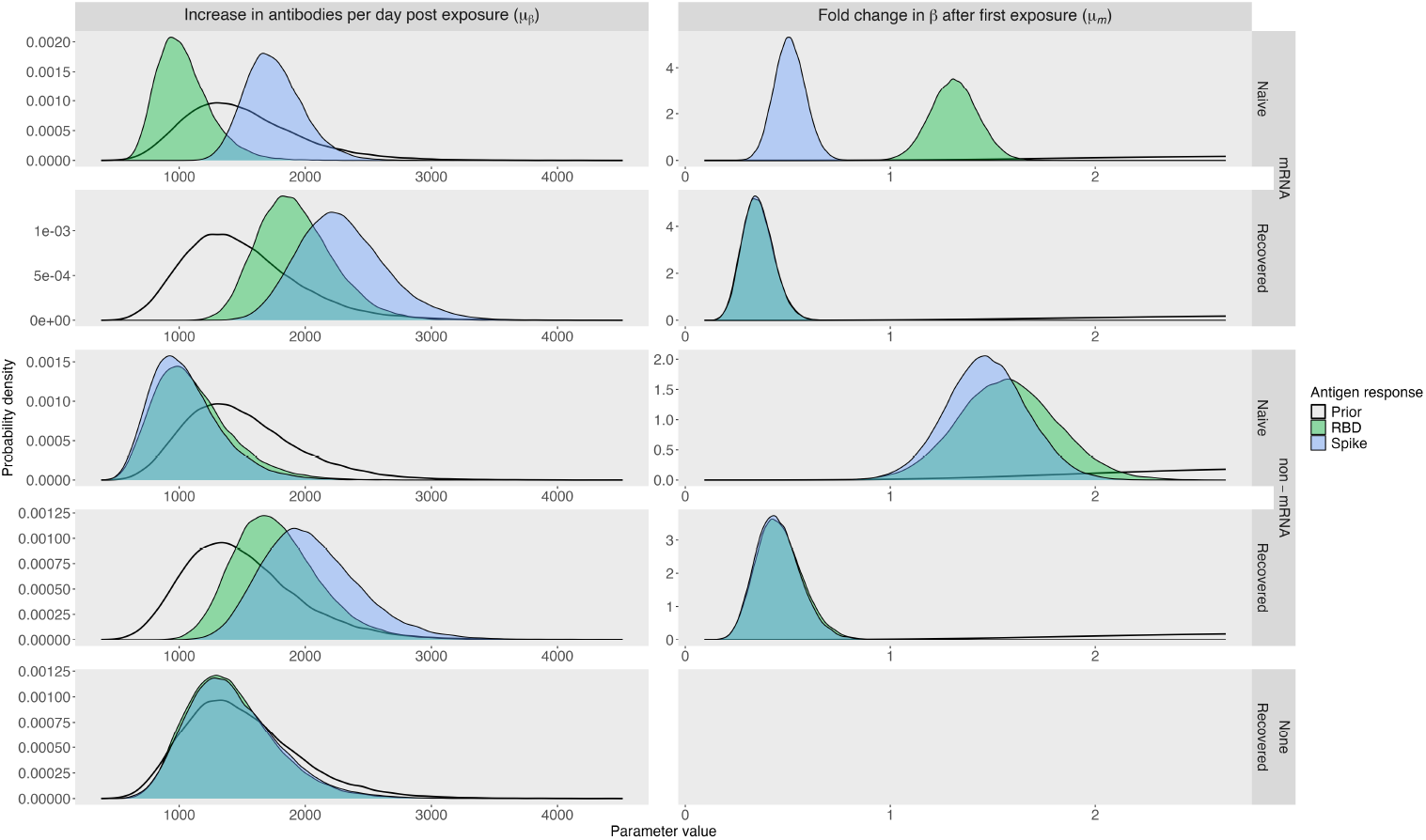
Population-level posterior parameter estimates relating to the boost in IgG antibodies upon exposure. The relevant parameters chosen were *μ*_*β*_, which represents the population-level increase in antibodies per day post exposure, and *μ*_*m*_, the population-level fold change in *β* after first exposure. The density plots represent the posterior distributions of the parameter estimates for the IgG response to RBD (green density plots), the IgG response to spike (blue density plots), and the prior distributions chosen (transparent density plot). Plots are organised by estimated obtained for each modelled group, separated by cohort and type of vaccination received, listed on the right. The x-axis represents the parameter values, being adjusted MFI for *μ*_*β*_, and fold change for *μ*_*m*_ and the y-axis is the probability density of the plots.

We then compared this to naive individuals who received mRNA and non-mRNA vaccinations. Those who received mRNA vaccinations were estimated to have a median increase in adjusted MFI of 1007.05 (95% CrI: [708.7, 1540.9]) for anti-RBD IgG, with a 17.2% probability of being higher than the estimates for those who experienced a natural infection alone. This was associated with a 1.31-fold increase (95% CrI: [1.09, 1.54]) in *β* for each exposure after the first vaccination. In contrast, anti-spike IgG antibodies were estimated to have a median increase of 1732.14 adjusted MFI (95% CrI: [1360.32, 2265.41]) in naive mRNA recipients, representing a 78.74% probability that the estimates were higher than those obtained for natural infection alone. We also observed a 0.51-fold reduction (95% CrI: [0.37, 0.66]) in the increase in anti-spike IgG following subsequent exposures after the first vaccination. These results indicate that, upon initial vaccination, the increase in anti-RBD antibodies is similar to that observed in natural infection alone; however, this increase becomes larger with subsequent vaccinations. Conversely, our results indicate that the production of anti-spike IgG is greater than that observed for natural infection alone; however, this production decreases after the first vaccination. The increase in anti-RBD IgG for naive individuals receiving non-mRNA vaccinations was estimated to be similar to that of naive mRNA recipients, with a median of 1060.18 (95% CrI: [638.28, 1854.16]) adjusted MFI. The antibody response to spike increased at a median of 1012.64 (95% CrI: [624.5, 1736.92]) adjusted MFI, with only a 19.87% probability that the estimates were greater than those obtained for natural infection alone. The naive non-mRNA vaccination recipients had a 1.47-fold (95% CrI: [1.09, 1.87]) and 1.57-fold (95% CrI: [1.11, 2.06]) increase in antibody production to the spike and RBD proteins, respectively, following the first vaccination. These results, similar to those for naive mRNA recipients, suggest that initial vaccination with a non-mRNA vaccine results in a similar increase in IgG titres as natural infection alone, which gets larger after the first vaccination.

Lastly, in Figure 2, we assess the increase in IgG for recovered individuals who also received vaccinations. Those who received mRNA vaccinations had a median increase of 2266.51 (95% CrI: [1701.58, 3063.62]) in adjusted MFI in response to spike protein, a 91.1% probability that the estimates were higher than those of their naive counterparts. Anti-RBD antibodies were estimated to increase with a median of 1898.26 (95% CrI: [1421.44, 2605.02]) adjusted MFI, indicating a probability of 99.14% that the estimates would be higher than those of naive individuals receiving mRNA vaccinations. Both anti-spike and anti-RBD IgG production were associated with an equal median fold-reduction of 0.35 (95% CrI: [0.22, 0.52] for spike, 95% CrI: [0.22, 0.52] for RBD) for further exposures, indicating a decrease in antibody production with vaccination following natural infection. Similarly, recovered individuals who received non-mRNA vaccinations had a 0.45-fold reduction (95% CrI: [0.27, 0.69]) and a 0.45-fold reduction (95% CrI: [0.27, 0.71]) in antibody production to spike and RBD antigens, respectively, across all vaccination types. These were also associated with a greater production of IgG upon exposure than naive counterparts in response to spike (median of 2011.68, 95% CrI: [1416.9, 2935.79]) and RBD (median of 1743.04, 95% CrI: [1211.24, 2581.65]). Full details on posterior population-level parameter estimates are provided in Table S5 and Table S6.

### 3.4 Half-life of IgG antibody responses to spike and RBD protein

#### 3.4.1 Results assuming a single exponential decay

Insight from the single exponential decay model reveals distinct differences from the biphasic exponential decay model in the half-life of the IgG response between the naive and recovered cohorts, as shown in Figure 3. When estimated through the single decay model, the median antibody response half-life for a natural infection alone was estimated as 318 days (95% CrI: [269.6, 386.8]) in response to spike and 343.4 (95% CrI: [285.4, 427.3]) to RBD, shown by the blue bars in the bottom panel of Figure 3. Estimates of IgG half-life for the spike antigen had a 99.16% probability of being higher for natural infection than those of the naive cohort who received mRNA vaccination (median half-life 220.1 days, 95% CrI: [178.7, 281.9]) and a 99.05% probability of being higher than the estimates for naive non-mRNA recipients (median half-life 210.7 days, 95% CrI: [166.4, 282.6]). Similar results are observed in response to RBD in Figure 3. These results suggest that the IgG response has a longer half-life after a single natural exposure to SARS-CoV-2 than after at least two doses of either an mRNA or non-mRNA vaccine. Recovered individuals who received mRNA vaccinations were estimated by the single phase decay model to have a median half-life of 452.6 days (95% CrI: 376.4, 556.3]) in response to spike, having a 100% probability that the estimates were higher than their naive counterparts, and a 99.42% probability of being higher than for that estimated for natural infection alone. Identical patterns are seen for recovered individuals receiving non-mRNA vaccinations and for the response to RBD protein (Table S4). This indicates that there was a substantial increase in the longevity of the IgG response in individuals who experienced a natural infection and received either an mRNA or non-mRNA vaccine schedule.

**Figure 3:**
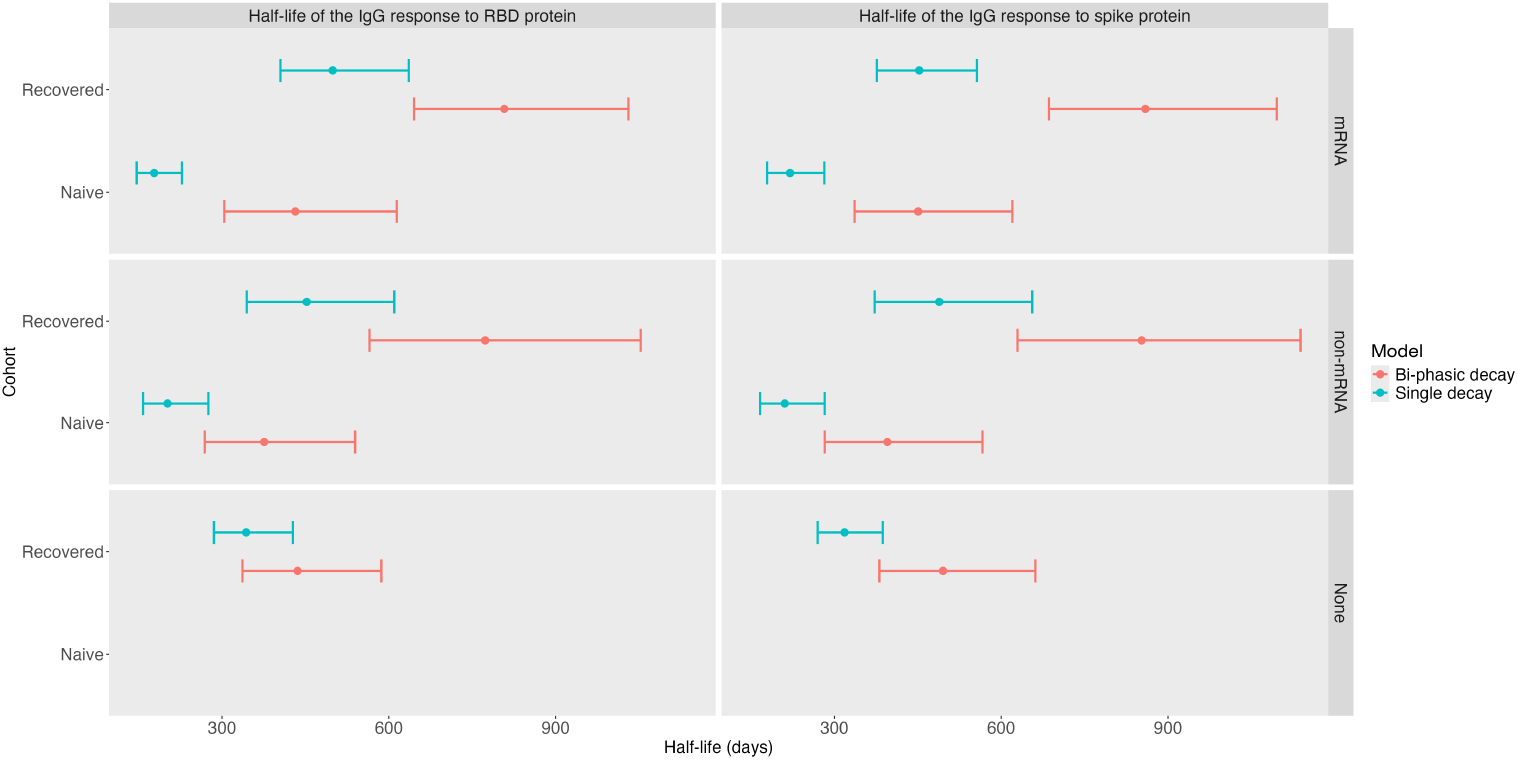
Population-level posterior parameter estimates for the IgG response half-life to spike and RBD antigens. Posterior parameter estimate medians are denoted by dots and 95% credible intervals are represented by the bars. Posterior estimates for 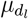 from the bi-phasic decay model and 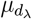 from the single phase decay model are shown in red and blue, respectively. The half-life of the IgG response is measured in days, represented on the x-axis, and estimates are separate by cohort (y-axis), type of vaccine received (right labels) and antigen response (top labels).

#### 3.4.2 Results assuming a bi-phasic exponential decay

Our interpretation of the half-life of the IgG response through the bi-phasic decay model is performed by estimating the half-life of the LLPCs 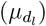.As seen in Figure 3, assuming antibodies decay in two phases leads to different results than the single-phase decay model estimations. The IgG response to spike protein for natural infection alone was estimated to have a median half-life of 495.2 days (95% CrI: 380.7, 661.2]) and 435.9 days (95% CrI: [336.6, 586.5]) in response to RBD, illustrated by the red bars in the bottom panel of Figure 3. The probability that estimates for the naive mRNA recipients were higher than for natural infection alone for spike protein was 32.54% (median half-life 450.6 days, 95% CrI: [336, 620]) and a 47.64% probability (median half-life 431.7 days, 95% CrI: [304.2, 614.2]) for RBD. Conversely, for naive non-mRNA recipients, there was a 15.79% probability (median half-life 395.2 days, 95% CrI: 282.6, 566.1]) of estimates being higher than the estimated longevity of the IgG response through natural infection alone to spike and a 25.53% probability (median half-life 375.9 days, 95% CrI: [268.8, 539.4]) for estimates for the response to RBD. These results suggest that the longevity of the IgG response elicited by mRNA vaccination has a higher probability of leading to a greater half-life compared to natural infection alone than when receiving non-mRNA vaccination.

A substantial difference is seen in Figure 3 for the recovered individuals who were vaccinated. The median half-life of the antibody response to spike protein was 859.2 days (95% CrI: [685.7, 1095.4]) for recovered mRNA recipients and 852.5 days (95% CrI:[629.4, 1138.4]) for non-mRNA recipients. These estimates have a 99.93% and 99.94% probability of being higher than those obtained for their mRNA and non-mRNA naive counterparts, respectively. Additionally, the longevity of the IgG response to spike antigen had a probability of being 99.828% higher in recovered mRNA recipients compared to estimates from natural infection alone and 99.44% for recovered non-mRNA recipients. The longevity of the anti-RBD IgG response demonstrates similar increases, with recovered mRNA recipients having a median half-life of 807.8 days (95% CrI: [645.4, 1030.9]) and non-mRNA recipients having a median half-life of 773.3 days ([565, 1053]). Similar to the results from the single-phase decay model, these estimates indicate a substantial increase in the longevity of the IgG response resulting from hybrid immunity in individuals who have experienced both a natural infection and vaccination.

Although the results are consistent across the models regarding which cohorts and vaccination types are estimated to have more extended IgG decay periods, the estimates between the two model types differ greatly. Both naive and recovered individuals receiving either mRNA or non-mRNA vaccination are estimated to have a median IgG response half-life, as determined by the bi-phasic decay model, almost twice that estimated by the single-phase decay model (Table S4-S6). Interestingly, the single exponential decay model estimated a longer antibody response half-life to RBD than spike for the recovered mRNA group and natural infection alone (Table S4). Conversely, the bi-phasic decay model estimated a higher median half-life for the IgG response to spike than RBD across all cohorts and vaccine groups (Table S5, Table S6). This suggests that the varying assumptions of both models can lead to different interpretations of the antibody response to the same antigen, resulting in conflicting outcomes. The posterior distributions for the population-level estimates for all model parameters are provided in Figure S2 and Figure S3.

### 3.5 Modelling IgG antibody kinetics to SARS-CoV-2 spike and RBD antigens

In Figure 4, we present the observed data and model fits for the IgG response to spike antigen for five representative individuals, one from each cohort-vaccination group that was modelled. We plot the fits from both the bi-phasic decay model and the single-phase decay model side by side for each individual, with the posterior predictive median and 95% posterior predictive interval calculated from IgG trajectory simulations generated from the posterior parameter estimates. Our results, as shown in Figure 4, visually demonstrate that the predicted IgG titres in response to exposure to the spike protein can accurately capture the boosting and decay through the bi-phasic model (left panel for each individual) and the decay through the single-phase model (right panel for each individual). We also notice that there are varying levels of uncertainty in the bi-phasic model results, as demonstrated by the width of the 95% posterior predictive interval.

**Figure 4:**
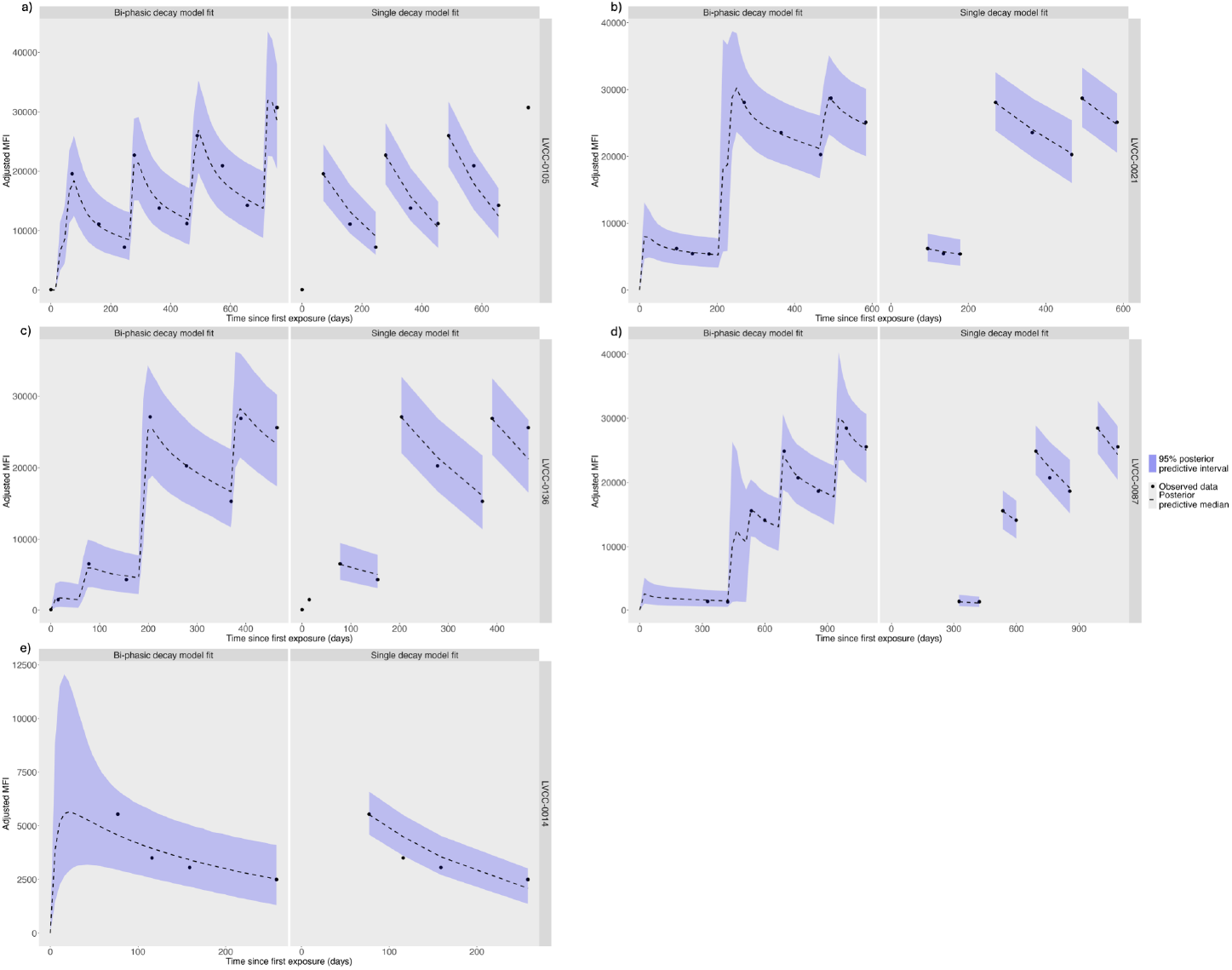
Bi-phasic decay model and single decay model fits with observed data for the IgG response to spike antigen for five representative individuals. Posterior predictive medians are represented by the dashed black line and the 95% posterior predictive median is shown by the blue ribbon. Observed data is indicated by black dots. **a)** Individual LVCC-0105 from the naive cohort that received mRNA vaccinations.**b)** Individual LVCC-0021 from the recovered cohort that received mRNA vaccinations. **c)** Individual LVCC-0136 from the naive cohort that received non-mRNA vaccinations. **d)** Individual LVCC-0087 from the recovered cohort that received non-mRNA vaccinations.**e)** Individual LVCC-0014 from the recovered cohort before receiving any vaccination.

There appears to be more uncertainty around the unobserved increases in antibodies, shown in Figures 4b and Figure 4d around the second exposure event, and in Figure 4e for the natural infection, given the increase in the posterior interval at these points. This is similarly seen with the IgG response to RBD in Figure 5, indicating that this uncertainty is reflective of missing observations and not due to the underlying kinetics of the IgG response.

**Figure 5:**
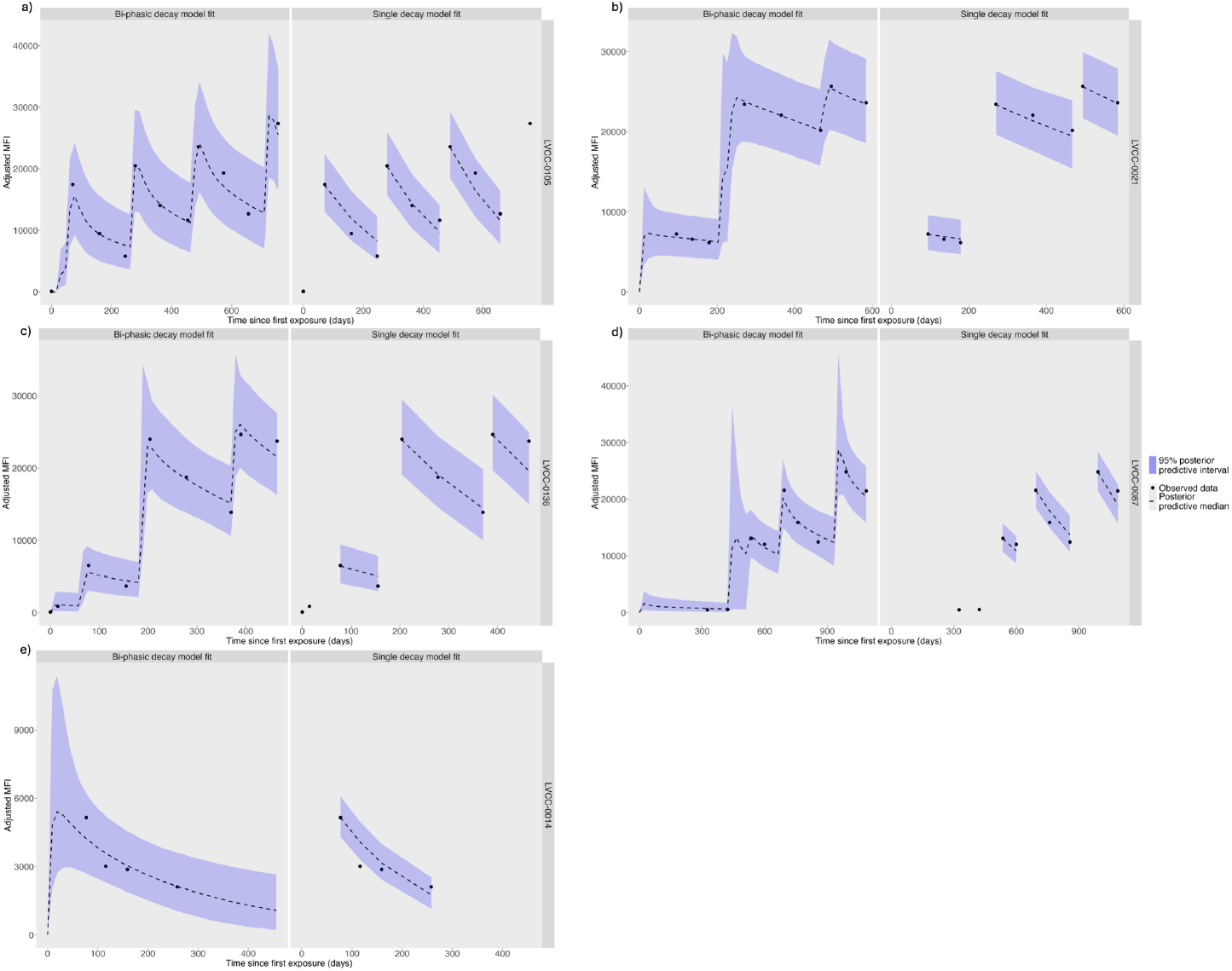
Bi-phasic decay model and single decay model fits with observed data for the IgG response to RBD antigen for five representative individuals. Posterior predictive medians are represented by the dashed black line and the 95% posterior predictive median is shown by the blue ribbon. Observed data is indicated by black dots. **a)** Individual LVCC-0105 from the naive cohort that received mRNA vaccinations.**b)** Individual LVCC-0021 from the recovered cohort that received mRNA vaccinations. **c)** Individual LVCC-0136 from the naive cohort that received non-mRNA vaccinations. **d)** Individual LVCC-0087 from the recovered cohort that received non-mRNA vaccinations.**e)** Individual LVCC-0014 from the recovered cohort before receiving any vaccination.

More information can be gathered visually about the antibody kinetics in response to the spike and RBD antigens through the bi-phasic exponential decay model than through the single exponential decay model. A notable piece of information missing from the plots of the single-phase decay model is the increasing portion of the IgG response. However, even focusing on the decay portion of the IgG response, we notice subtle differences. There is a change in the decline of the posterior predictive median line as the short-lived phase of the antibody response ends and the long-lived maintenance phase mediated by long-lived plasma cells takes over, indicated by the line plateauing. This is particularly evident in Figure 4b and Figure 4c during the IgG decay after the second exposure event. Almost identical results are seen when plotting the model fits for the same individuals’ IgG response to RBD (Figure 5). These results indicate that estimates gathered through the bi-phasic decay model are more informative about the underlying mechanisms behind IgG production in response to the spike and RBD than those obtained from the single-phase model. Complete antibody trajectories generated through posterior parameter estimates from the bi-phasic exponential decay model for all individuals are provided in Figures S4-S13 and for the single exponential decay model in Figures S14-S23.

## 4. Discussion

Despite the extensive number of studies published on humoral immunity to SARS-CoV-2, there remains ambiguity regarding the duration of the antibody response following infection. Given this, we investigated the antibody kinetics across individuals exposed to SARS-CoV-2 through natural infection and vaccination to quantify the IgG response half-life and identify the differences between hybrid immunity and the humoral immunity conferred by either vaccination or infection alone. We estimated the IgG response halflife using a simple exponential model, assuming that antibody decay is constant, and a bi-phasic exponential decay model, which assumes a short- and long-lived component of IgG waning and allows us to estimate the strength of the IgG response upon exposure. This was done not only to illustrate the varying results that different model assumptions can yield but also to determine if patterns across exposure groups (e.g., a specific type of vaccine eliciting a stronger response) were consistent across different assumptions. The results in this work demonstrate that after a full primary-vaccination schedule (two doses), recipients of an mRNA vaccination had significantly higher IgG levels than recipients of a non-mRNA vaccination. This is in line with previous studies that have found that the BNT162b2 vaccine is more effective in generating a greater antibody response than the ChAdOx1-S vaccine [17, 18, 38, 60]. We also found that there was no significant change in the anti-spike or anti-RBD IgG levels of the recovered cohort between the first and second vaccination, regardless of the type of vaccination received, while there was in the naive cohort. This has been identified in previously infected individuals who received the BNT162b2 vaccine [18, 61, 62], but our results demonstrate that it also applies to those receiving other non-mRNA vaccines.

Further confirming our results, studies have shown that only one vaccination is required for recently infected individuals to achieve their highest antibody level [18, 62, 63], which may explain why there is no further increase in the maximum IgG titre after the first vaccination for the recovered cohort. Interestingly, the most significant increase in both the spike-specific and RBD-specific IgG titres observed in the COVID PROFILE data was in naive non-mRNA recipients between their second dose and their booster vaccination. This could be the result of heterologous vaccination, as all participants in the COVID PROFILE study who received ChAdOx1-S for their first two doses subsequently received an mRNA vaccination (the majority received BNT162b2) for their booster. These results are in line with studies that have found a greater increase in SARS-CoV-2 specific IgG antibodies after heterologous vaccination with an initial two-dose vaccine schedule of a non-mRNA vaccine, followed by a booster mRNA vaccine [17, 63–66].

Although individuals from the recovered cohort demonstrated consistently high spike-specific and RBD-specific antibody titres, examination of the underlying IgG kinetics shows a reduction in the increase of antibodies produced with additional exposure. This is in line with results from other studies which show a reduced IgG response in previously infected individuals after vaccination and is suggested to be due to the presence of a higher level of antibodies pre-exposure compared to naive individuals [18, 39, 54, 67, 68]. Our results indicate that the reason that recovered individuals maintain higher IgG titres than naive individuals is related to a longer antibody response half-life observed with hybrid immunity. This would also explain why naive individuals exhibit a larger increase in anti-spike and anti-RBD IgG production after exposure. It is not necessarily reflective of a stronger antibody response but rather a lower baseline of IgG levels before vaccination, given that the estimated half-life of the IgG response in the naive cohort is half the duration of those with hybrid immunity. As these results are consistent across bi-phasic decay modelling and single exponential decay modelling of the IgG response to both spike and RBD, we suggest that they reflect changes in immunity that enhance the long-term maintenance of the humoral response elicited only through hybrid immunity, which is not obtained through natural infection or vaccination alone. Others have alluded to this [7, 14–16, 69], but it was previously unquantified in the literature until this study. Importantly, it had not previously been quantified accounting for the boosting component of the antibody response and assuming bi-phasic exponential decay.

Our posterior parameter estimates from both models demonstrate the impact that different model assumptions can have on results, as we found drastically different results depending on our assumptions about antibody decay. This has been observed in other studies modelling the antibody response to spike, RBD, and nucleocapsid [35, 36]. Grandjean et al. (2022) [36] observed that the difference in assuming single-phase decay or bi-phasic decay through different gamma models did not produce different results for the short-lived IgG response to nucleocapsid or RBD. However, a significant change was observed in the estimation of the antibody response half-life to spike protein. However, Cohen et al. (2021) [35] found that a power law model, accounting for bi-phasic decay, was better suited to their data and led to greater half-life estimates for the IgG response to nucleocapsid, RBD, and spike than that of single exponential decay. This has important implications for our conclusions regarding long-term immunity to SARS-CoV-2. Given that a single-phase decay model neglects differences in short- and long-lived dynamics, it can underestimate the longevity of the antibody response and impact decisions on vaccine schedules and cost-effectiveness analysis, which are based on information on antibody waning [70]. Underestimating the half-life of the antibody response can result in vaccine schedules with shorter intervals, which can produce ineffective antibody responses [71]. This has important implications not only for SARS-CoV-2 but also for any other existing or novel pathogens that may arise in the future, for which we need appropriate vaccination schedules. Additionally, the type of antibody kinetics model we use can assess the impact and longevity of the antibody response to vaccination differently and have consequences for the type and number of vaccines recommended for different cohorts of people. These all have downstream consequences for public health decisions and the health of the population. Thus, we highlight through our analyses using different models, the importance of checking model assumptions and how they align with biological mechanisms we know to be true in response to infectious diseases. Given that we understand the antibody response to SARS-CoV-2 to have a bi-phasic wane [72–74], we find the results from our bi-phasic exponential decay model, rather than the single exponential decay, to be of importance.

There are several limitations to this analysis. Although the bi-phasic exponential decay model is better suited to the data than the single exponential decay model, it still has its own limiting assumptions. The model does not consider any delay in the initiation of the humoral immune response after primary exposure to SARS-CoV-2 and does not account for changes in the proportion of short-lived plasma cell population over time, which we know to occur during infection [75–78]. Additionally, due to identifiability issues with model parameters, we are unable to estimate the short-lived and long-lived plasma cell populations, as well as the antibody secretion rate. Based on this, we cannot determine how much variation in the antibody response is attributable to each of these parameters. The model also does not account for changes in the half-life of long-lived plasma cells (LLPCs) over time for an individual, instead assuming that it remains constant. Given our data and the short interval between vaccinations, it would be challenging to determine if there is a change in LLPC half-life throughout individual follow-up. This leads to a further limitation of our data. For our groups of individuals with hybrid immunity (recovered individuals who have received either mRNA or non-mRNA vaccination), we do not know how many vaccinations are required to significantly increase the IgG response half-life compared to the naive cohort or natural infection alone. A study involving individuals who have previously been infected and only received one, two, or three vaccinations could provide greater insight into the impact of the number of vaccines received on the longevity of the antibody response in recovered individuals. Additionally, this information could provide a way to measure changes in the proportion of SLPCs and the half-life of LLPCs across multiple exposures. The time between natural infection and the first blood sample collection for the recovered cohort, which for many is their antibody production phase and initial antibody decay, is unobserved in our data and may have led to parameter estimates relying more heavily on prior information due to the limited number of samples available to inform them. A study that collects blood samples in the initial weeks after vaccination could provide this data and lead to more informed estimates.

Another limitation of our data is that serology was based on ancestral antigens of SARS-CoV-2. Throughout the time frame of the COVID PROFILE study, circulating viruses and vaccines changed from the ancestral strain. To account for infection with different viral strains, we removed any observations for individuals who were naturally infected during the study. However, some individuals enrolled in the study continued to receive vaccinations until April 4, 2023, and could have received one of the bivalent vaccines provisionally approved in late 2022 and early 2023 by the Therapeutic Goods Administration of Australia, which contained ancestral and Omicron strains of SARS-CoV-2 [79]. We suggest that the bivalent vaccines would have a similar impact on the boosting and maintenance of IgG antibodies against the ancestral spike and the RBD antigen as the monovalent ancestral vaccines. Firstly, the ancestral spike antigen was included in the Moderna and Pfizer bivalent vaccines, which would stimulate immune recall in individuals previously infected and/or vaccinated with the monovalent vaccine and lead to more antibody production [34, 80, 81]. Second, there is strong evidence in the literature for the presence of immune imprinting for SARS-CoV-2 [69, 82–85], suggesting that individuals primed by the ancestral strain would be back-boosted by exposure to variant antigens. We find no substantial evidence in the literature to suggest a significantly increased or decreased ancestral spike-specific and RBD-specific IgG response resulting from vaccination with the bivalent ancestral and Omicron strain vaccines compared to the antibody response from the first monovalent SARS-CoV-2 mRNA vaccines.

A notable consideration is that the COVID PROFILE data does not include serology of neutralising antibodies, which are recognised as a proxy correlate of immune protection. Our analysis was focused only on total IgG binding antibodies. Neutralising antibodies have important relevance for protection against SARS-CoV-2 variants of concern, with a high titre of said antibodies being associated with a decreased risk of reinfection and severe disease [67, 86–88]. A strong correlation has been noted between spike-specific binding IgG antibody titres and neutralising antibody levels [35, 89–94], indicating that measuring binding IgG antibodies can provide some information about immune protection. Therefore, we reason that our work is important when considering protective immunity. There is no established protective threshold that can be used with serological testing to define protection against reinfection [88]. However, it is well understood that high titres of neutralising antibodies result in greater protection. Thus, a high level of binding IgG can also indicate greater protection. This may be demonstrated in our results, where we have shown that a single vaccination is suffcient for SARS-CoV-2 recovered individuals to reach their highest IgG antibody titre, and other studies have shown greater protection against variants of concern for those with hybrid immunity [7, 12, 14–16]. As we have also shown that hybrid immunity results in a longer half-life of the IgG response, this may suggest that immune protection also lasts longer for recovered individuals after vaccination. The WHO has recommended a single dose of an mRNA vaccine every 12 months for previously infected individuals to maintain population-level protective immunity, given their high antibody titres and sustained response after vaccination [71], in line with our results from the bi-phasic exponential decay model. Thus, we demonstrate that our results from antibody kinetics modelling on binding IgG antibodies can provide important information on the maintenance of protective immunity, as do studies on neutralising antibodies. This can be a helpful alternative given that assays to measure binding antibodies have more reproducible results, lower costs, and are less complex than neutralisation assays [94].

Additionally, this has important implications for informing public health decisions on modifying vaccine regimes to suit populations with specific immune profiles. This application extends beyond the case study of SARS-CoV-2. We emphasise that modelling of antibody production and decay is of great use to better understand immunity at both an individual- and population-level, to determine appropriate vaccine scheduling [26, 27, 95, 96], and for identification of clinical and subclinical cases of disease [97] for a range of different pathogens.

## 5. Conclusions

A comprehensive understanding of long-term humoral immunity to SARS-CoV-2 is crucial for identifying high-risk populations and making informed public health decisions to maintain population-level protection. Our findings show that hybrid immunity extends the longevity of the IgG response to RBD and spike protein by almost double that obtained through vaccination or natural infection alone. Additionally, we demonstrate that a single dose of either an mRNA or non-mRNA vaccine is suffcient for previously infected individuals to regain peak antibody titres. By using two separate models to analyse our data, we show the impact that different assumptions can have on the estimation of IgG decay half-life and discuss the importance of considering these when using model results to inform decisions. Our study provides a framework that can assist in assessing the longevity of the humoral response, thereby better preparing for future pandemics and the emergence of novel viruses. This framework can help determine population-level immunity and could be used to inform long-term recommendations for vaccine scheduling and booster vaccine scenarios.

## Data Availability

Data is available upon reasonable request to Emily Eriksson (email address).

## References

1. Gazit, S. et al. Severe Acute Respiratory Syndrome Coronavirus 2 (SARS-CoV-2) Naturally Acquired Immunity versus Vaccine-induced Immunity, Reinfections versus Breakthrough Infections: A Retrospective Cohort Study. Clinical Infectious Diseases 75, e545–e551. ISSN: 1058-4838. 10.1093/cid/ciac262 (2024) (July 2022).

2. Wendelboe, A. M., Van Rie, A., Salmaso, S., & Englund, J. A. Duration of Immunity Against Pertussis After Natural Infection or Vaccination. en-US. The Pediatric Infectious Disease Journal 24, S58. ISSN: 0891-3668. https://journals.lww.com/pidj/fulltext/2005/05001/duration_of_immunity_against_pertussis_after.11.as (2024) (May 2005).

3. Zimmermann, P. & Curtis, N. Factors That Influence the Immune Response to Vaccination. EN. Clinical Microbiology Reviews. Publisher: American Society for Microbiology1752 N St., N.W., Washington, DC. https://journals.asm.org/doi/10.1128/cmr.00084-18 (2024) (Mar. 2019).

4. Bianchi, F. P. et al. Long-term immunogenicity after measles vaccine vs. wild infection: an Italian retrospective cohort study. Human Vaccines & Immunotherapeutics 17, 2078–2084. ISSN: 2164-5515. https://www.ncbi.nlm.nih.gov/pmc/articles/PMC8189124/ (2024) (Jan. 2021).

5. Pooley, N. et al. Durability of Vaccine-Induced and Natural Immunity Against COVID-19: A Narrative Review. en. Infectious Diseases and Therapy 12, 367–387. ISSN: 2193-6382. 10.1007/s40121-022-00753-2 (2025) (Feb. 2023).

6. Pérez-Alós, L. et al. Modeling of waning immunity after SARS-CoV-2 vaccination and influencing factors. en. Nature Communications 13. Publisher: Nature Publishing Group, 1614. ISSN: 2041-1723. https://www.nature.com/articles/s41467-022-29225-4 (2025) (Mar. 2022).

7. Goldberg, Y. et al. Protection and Waning of Natural and Hybrid Immunity to SARS-CoV-2. New England Journal of Medicine 386. Publisher: Massachusetts Medical Society, 2201–2212. ISSN: 0028-4793. https://www.nejm.org/doi/full/10.1056/NEJMoa2118946 (2024) (June 2022).

8. Chemaitelly, H. et al. Protection from previous natural infection compared with mRNA vaccination against SARS-CoV-2 infection and severe COVID-19 in Qatar: a retrospective cohort study. The Lancet Microbe 3. Publisher: Elsevier, e944–e955 (2022).

9. Franchi, M. et al. Natural and vaccine-induced immunity are equivalent for the protection against SARS-CoV-2 infection. Journal of Infection and Public Health 16, 1137–1141. ISSN: 1876-0341. https://www.sciencedirect.com/science/article/pii/S1876034123001739 (2024) (Aug. 2023).

10. Lumley, S. F. et al. An Observational Cohort Study on the Incidence of Severe Acute Respiratory Syndrome Coronavirus 2 (SARS-CoV-2) Infection and B.1.1.7 Variant Infection in Healthcare Workers by Antibody and Vaccination Status. en. Clinical Infectious Diseases 74, 1208–1219. ISSN: 1058-4838, 1537-6591. https://academic.oup.com/cid/article/74/7/1208/6314286 (2024) (Apr. 2022).

11. Goldberg, Y. et al. Similarity of Protection Conferred by Previous SARS-CoV-2 Infection and by BNT162b2 Vaccine: A 3-Month Nationwide Experience From Israel. American Journal of Epidemiology 191, 1420–1428. ISSN: 0002-9262. 10.1093/aje/kwac060 (2024) (July 2022).

12. Shenai, M. B., Rahme, R., & Noorchashm, H. Equivalency of Protection From Natural Immunity in COVID-19 Recovered Versus Fully Vaccinated Persons: A Systematic Review and Pooled Analysis. Cureus 13, e19102. ISSN: 2168-8184. https://www.ncbi.nlm.nih.gov/pmc/articles/PMC8627252/ (2025) (Oct. 2021).

13. Ibarrondo, F. J. et al. Primary, Recall, and Decay Kinetics of SARS-CoV-2 Vaccine Antibody Responses. ACS Nano 15. Publisher: American Chemical Society, 11180–11191. ISSN: 1936-0851. 10.1021/acsnano.1c03972 (2025) (July 2021).

14. Hall, V. et al. Protection against SARS-CoV-2 after Covid-19 Vaccination and Previous Infection. New England Journal of Medicine 386. Publisher: Massachusetts Medical Society, 1207–1220. ISSN: 0028-4793. https://www.nejm.org/doi/full/10.1056/NEJMoa2118691 (2025) (Mar. 2022).

15. Hammerman, A. et al. Effectiveness of the BNT162b2 Vaccine after Recovery from Covid-19. New England Journal of Medicine 386. Publisher: Massachusetts Medical Society, 1221–1229. ISSN: 0028-4793. https://www.nejm.org/doi/full/10.1056/NEJMoa2119497 (2025) (Mar. 2022).

16. Spinardi, J. R. & Srivastava, A. Hybrid Immunity to SARS-CoV-2 from Infection and Vaccination—Evidence Synthesis and Implications for New COVID-19 Vaccines. en. Biomedicines 11. Number: 2 Publisher: Multidisciplinary Digital Publishing Institute, 370. ISSN: 2227-9059. https://www.mdpi.com/2227-9059/11/2/370 (2024) (Feb. 2023).

17. Rose, R. et al. Humoral immune response after different SARS-CoV-2 vaccination regimens. BMC Medicine 20, 31. ISSN: 1741-7015. 10.1186/s12916-021-02231-x (2025) (Jan. 2022).

18. Goel, R. R. et al. Distinct antibody and memory B cell responses in SARS-CoV-2 naïve and recovered individuals after mRNA vaccination. Science Immunology 6. Publisher: American Association for the Advancement of Science, eabi6950. https://www.science.org/doi/10.1126/sciimmunol.abi6950 (2025) (Apr. 2021).

19. Abd-alrazaq, A. et al. Machine Learning–Based Approach for Identifying Research Gaps: COVID-19 as a Case Study. EN. JMIR Formative Research 8. Publisher: JMIR Publications Inc., Toronto, Canada, e49411. https://formative.jmir.org/2024/1/e49411 (2024) (Mar. 2024).

20. Burton, D. R. Antibodies, viruses and vaccines. en. Nature Reviews Immunology 2. Publisher: Nature Publishing Group, 706–713. ISSN: 1474-1741. https://www.nature.com/articles/nri891 (2024) (Sept. 2002).

21. Zohar, T. & Alter, G. Dissecting antibody-mediated protection against SARS-CoV-2. en. Nature Reviews Immunology 20. Publisher: Nature Publishing Group, 392–394. ISSN: 1474-1741. https://www.nature.com/articles/s41577-020-0359-5 (2024) (July 2020).

22. Padilla-Quirarte, H. O., Lopez-Guerrero, D. V., Gutierrez-Xicotencatl, L., & Esquivel-Guadarrama, F. Protective Antibodies Against Influenza Proteins. English. Frontiers in Immunology 10.Publisher: Frontiers. ISSN: 1664-3224. https://www.frontiersin.org/journals/immunology/articles/10.3389/fimmu.2019.01677/full (2024) (July 2019).

23. Forthal, D. N. Functions of Antibodies. Microbiology Spectrum 2. Publisher: American Society for Microbiology, 10.1128/microbiolspec.aid–0019–2014. https://journals.asm.org/doi/full/10.1128/microbiolspec.aid-0019-2014 (2024) (Aug. 2014).

24. Slifka, M. K. & Ahmed, R. Long-lived plasma cells: a mechanism for maintaining persistent antibody production. Current Opinion in Immunology 10, 252–258. ISSN: 0952-7915. https://www.sciencedirect.com/science/article/pii/S0952791598801623 (2025) (June 1998).

25. White, M. T. et al. Dynamics of the Antibody Response to Plasmodium falciparum Infection in African Children. The Journal of Infectious Diseases 210, 1115–1122. ISSN: 0022-1899. 10.1093/infdis/jiu219 (2023) (Oct. 2014).

26. Andraud, M. et al. Living on Three Time Scales: The Dynamics of Plasma Cell and Antibody Populations Illustrated for Hepatitis A Virus. en. PLOS Computational Biology 8. Publisher: Public Library of Science, e1002418. ISSN: 1553-7358. https://journals.plos.org/ploscompbiol/article?id=10.1371/journal.pcbi.1002418 (2025) (Mar. 2012).

27. Le, D., Miller, J. D., & Ganusov, V. V. Mathematical modeling provides kinetic details of the human immune response to vaccination. Frontiers in Cellular and Infection Microbiology 4. ISSN: 2235-2988. https://www.frontiersin.org/articles/10.3389/fcimb.2014.00177 (2023) (2015).

28. Van Elslande, J., Gruwier, L., Godderis, L., & Vermeersch, P. Estimated Half-Life of SARS-CoV-2 Anti-Spike Antibodies More Than Double the Half-Life of Antinucleocapsid Antibodies in Healthcare Workers. Clinical Infectious Diseases 73, 2366–2368. ISSN: 1058-4838. https://doi.org/10.1093/cid/ciab219 (2024) (Dec. 2021).

29. Vanshylla, K. et al. Kinetics and correlates of the neutralizing antibody response to SARS-CoV-2 infection in humans. en. Cell Host & Microbe 29, 917–929.e4. ISSN: 19313128. https://linkinghub.elsevier.com/retrieve/pii/S1931312821001918 (2024) (June 2021).

30. Cimas, F. J. et al. Mathematical modelling of the waning of anti-RBD IgG SARS-CoV-2 antibody titers after a two-dose BNT162b2 mRNA vaccination. Frontiers in Immunology 14. ISSN: 1664-3224. https://www.frontiersin.org/articles/10.3389/fimmu.2023.1097747 (2023) (2023).

31. Whitcombe, A. L. et al. Comprehensive analysis of SARS-CoV-2 antibody dynamics in New Zealand. Clinical & Translational Immunology 10, e1261. ISSN: 2050-0068. https://www.ncbi.nlm.nih.gov/pmc/articles/PMC7955949/ (2025) (Mar. 2021).

32. Yorsaeng, R. et al. SARS-CoV-2 Antibody Dynamics after COVID-19 Vaccination and Infection: A Real-World Cross-Sectional Analysis. en. Vaccines 11. Number: 7 Publisher: Multidisciplinary Digital Publishing Institute, 1184. ISSN: 2076-393X. https://www.mdpi.com/2076-393X/11/7/1184 (2025) (July 2023).

33. Lumley, S. F. et al. The Duration, Dynamics, and Determinants of Severe Acute Respiratory Syndrome Coronavirus 2 (SARS-CoV-2) Antibody Responses in Individual Healthcare Workers. Clinical Infectious Diseases: An Offcial Publication of the Infectious Diseases Society of America 73, e699–e709. ISSN: 1058-4838. https://www.ncbi.nlm.nih.gov/pmc/articles/PMC7929225/ (2025) (Jan. 2021).

34. Chen, Y. et al. Immune recall improves antibody durability and breadth to SARS-CoV-2 variants. Science Immunology 7. Publisher: American Association for the Advancement of Science, eabp8328. https://www.science.org/doi/full/10.1126/sciimmunol.abp8328 (2025) (May 2022).

35. Cohen, K. W. et al. Longitudinal analysis shows durable and broad immune memory after SARS-CoV-2 infection with persisting antibody responses and memory B and T cells. en. Cell Reports Medicine 2, 100354. ISSN: 26663791. https://linkinghub.elsevier.com/retrieve/pii/S2666379121002032 (2024) (July 2021).

36. Grandjean, L. et al. Long-Term Persistence of Spike Protein Antibody and Predictive Modeling of Antibody Dynamics After Infection With Severe Acute Respiratory Syndrome Coronavirus 2. Clinical Infectious Diseases 74, 1220–1229. ISSN: 1058-4838. 10.1093/cid/ciab607 (2024) (Apr. 2022).

37. Rosado, J. et al. Multiplex assays for the identification of serological signatures of SARS-CoV-2 infection: an antibody-based diagnostic and machine learning study. en. The Lancet Microbe 2, e60–e69. ISSN: 2666-5247. https://www.sciencedirect.com/science/article/pii/S266652472030197X (2023) (Feb. 2021).

38. Stocks, D., Thomas, A., Finn, A., Danon, L., & Brooks-Pollock, E. Mechanistic models of humoral kinetics following COVID-19 vaccination. Journal of The Royal Society Interface 22. Publisher: Royal Society, 20240445. https://royalsocietypublishing.org/doi/full/10.1098/rsif.2024.0445 (2025) (Jan. 2025).

39. Nishiyama, T. et al. Modeling COVID-19 vaccine booster-elicited antibody response and impact of infection history. Vaccine 41, 7655–7662. ISSN: 0264-410X. https://www.sciencedirect.com/science/article/pii/S0264410×23013841 (2025)(Dec. 2023).

40. Deichmann, J. et al. Predicting antibody kinetics and duration of protection against SARS-CoV-2 following vaccination from sparse serological data. en. PLOS Computational Biology 21. Publisher: Public Library of Science, e1013192. ISSN: 1553-7358. https://journals.plos.org/ploscompbiol/article?id=10.1371/journal.pcbi.1013192 (2025) (June 2025).

41. Eriksson, E. M. et al. Cohort Profile: A longitudinal Victorian COVID-19 cohort (COVID PROFILE) en. preprint (Infectious Diseases (except HIV/AIDS), Apr. 2023). http://medrxiv.org/lookup/doi/10.1101/2023.04.27.23289157 (2024).

42. Mazhari, R. et al. SARS-CoV-2 Multi-Antigen Serology Assay. en. Methods and Protocols 4. Number: 4 Publisher: Multidisciplinary Digital Publishing Institute, 72. ISSN: 2409-9279. https://www.mdpi.com/2409-9279/4/4/72 (2024) (Dec. 2021).

43. White, M. et al. Antibody kinetics following vaccination with MenAfriVac: an analysis of serological data from randomised trials. en. The Lancet Infectious Diseases 19, 327–336. ISSN: 1473-3099. https://www.sciencedirect.com/science/article/pii/S1473309918306741 (2023) (Mar. 2019).

44. Liu, Z. S.-J. et al. Naturally acquired antibody kinetics against Plasmodium vivax antigens in people from a low malaria transmission region in western Thailand. BMC Medicine 20, 89. ISSN: 1741-7015. 10.1186/s12916-022-02281-9 (2023) (Mar. 2022).

45. Hammarlund, E. et al. Plasma cell survival in the absence of B cell memory. en. Nature Communications 8. Publisher: Nature Publishing Group, 1781. ISSN: 2041-1723. https://www.nature.com/articles/s41467-017-01901-w (2025) (Nov. 2017).

46. Wilmore, J. R. & Allman, D. Here, There, and Anywhere? Arguments for and against the Physical Plasma Cell Survival Niche. The Journal of Immunology 199, 839–845. ISSN: 0022-1767. 10.4049/jimmunol.1700461 (2025) (Aug. 2017).

47. Moser, K. et al. Long-lived plasma cells in immunity and immunopathology. Immunology Letters 103, 83–85. ISSN: 0165-2478. https://www.sciencedirect.com/science/article/pii/S0165247805002919 (2025) (Mar. 2006).

48. Cocco, M. et al. In Vitro Generation of Long-lived Human Plasma Cells. The Journal of Immunology 189, 5773–5785. ISSN: 0022-1767. 10.4049/jimmunol.1103720 (2025) (Dec. 2012).

49. Foss, S. et al. Human IgG Fc-engineering for enhanced plasma half-life, mucosal distribution and killing of cancer cells and bacteria. en. Nature Communications 15. Publisher: Nature Publishing Group, 2007. ISSN: 2041-1723. https://www.nature.com/articles/s41467-024-46321-9 (2025) (Mar. 2024).

50. Saxena, A. & Wu, D. Advances in Therapeutic Fc Engineering – Modulation of IgG-Associated Effector Functions and Serum Half-life. English. Frontiers in Immunology 7. Publisher: Frontiers. ISSN: 1664-3224. https://www.frontiersin.org/journals/immunology/articles/10.3389/fimmu.2016.00580/full (2025) (Dec. 2016).

51. Morell, A., Terry, W. D., & Waldmann, T. A. Metabolic properties of IgG subclasses in man. en. The Journal of Clinical Investigation 49. Publisher: American Society for Clinical Investigation, 673–680. ISSN: 0021-9738. https://www.jci.org/articles/view/106279 (2025) (Apr. 1970).

52. Grevys, A. et al. Antibody variable sequences have a pronounced effect on cellular transport and plasma half-life. iScience 25, 103746. ISSN: 2589-0042. https://www.sciencedirect.com/science/article/pii/S2589004222000165 (2025) (Feb. 2022).

53. Stan Development Team. Prior Choice Recommendations en. Mar. 2024. https://github.com/stan-dev/stan/wiki/Prior-Choice-Recommendations (2025).

54. Misra, A. & Theel, E. S. Immunity to SARS-CoV-2: What Do We Know and Should We Be Testing for It? Journal of Clinical Microbiology 60. Publisher: American Society for Microbiology, e00482–21. https://journals.asm.org/doi/10.1128/jcm.00482-21 (2025) (Mar. 2022).

55. Carpenter, B. et al. Stan: A Probabilistic Programming Language. Journal of statistical software 76, 1. ISSN: 1548-7660. https://www.ncbi.nlm.nih.gov/pmc/articles/PMC9788645/ (2023) (2017).

56. Hoffman, M. D. & Gelman, A. The No-U-Turn Sampler: Adaptively Setting Path Lengths in Hamiltonian Monte Carlo. en. Journal of Machine Learning Research 15, 1593–1623 (2014).

57. Neal, R. M. in Handbook of Markov Chain Monte Carlo Num Pages: 50 (Chapman and Hall/CRC, 2011). isbn: 978-0-429-13850-8.

58. Stan Development Team. RStan: the R interface to Stan 2024. https://mc-stan.org/.

59. R Core Team. R: A Language and Environment for Statistical Computing https://www.R-project.org/ (R Foundation for Statistical Computing, Vienna, Austria, 2023).

60. Reynolds, L., Dewey, C., Asfour, G., & Little, M. Vaccine efficacy against SARS-CoV-2 for Pfizer BioNTech, Moderna, and AstraZeneca vaccines: a systematic review. English. Frontiers in Public Health 11. Publisher: Frontiers. ISSN: 2296-2565. https://www.frontiersin.org/journals/public-health/articles/10.3389/fpubh.2023.1229716/full (2025) (Oct. 2023).

61. Ebinger, J. E. et al. Antibody responses to the BNT162b2 mRNA vaccine in individuals previously infected with SARS-CoV-2. en. Nature Medicine 27. Publisher: Nature Publishing Group, 981–984. ISSN: 1546-170X. https://www.nature.com/articles/s41591-021-01325-6 (2025) (June 2021).

62. Krammer, F. et al. Antibody Responses in Seropositive Persons after a Single Dose of SARS-CoV-2 mRNA Vaccine. New England Journal of Medicine 384. Publisher: Massachusetts Medical Society, 1372–1374. ISSN: 0028-4793. https://www.nejm.org/doi/full/10.1056/NEJMc2101667 (2025) (Apr. 2021).

63. Moore, S. C. et al. Evolution of long-term vaccine-induced and hybrid immunity in healthcare workers after different COVID-19 vaccine regimens. English. Med 4. Publisher: Elsevier, 191–215.e9. ISSN: 2666-6359, 2666-6340. https://www.cell.com/med/abstract/S2666-6340(23)00064-8 (2024) (Mar. 2023).

64. Ho, T.-C. et al. The Effects of Heterologous Immunization with Prime-Boost COVID-19 Vaccination against SARS-CoV-2. en. Vaccines 9. Number: 10 Publisher: Multidisciplinary Digital Publishing Institute, 1163. ISSN: 2076-393X. https://www.mdpi.com/2076-393X/9/10/1163 (2025) (Oct. 2021).

65. Zhang, J. et al. Boosting with heterologous vaccines effectively improves protective immune responses of the inactivated SARS-CoV-2 vaccine. Emerging Microbes & Infections 10, 1598–1608. ISSN: 2222-1751. https://www.ncbi.nlm.nih.gov/pmc/articles/PMC8381941/ (2025) (Aug. 2021).

66. Zhang, X. et al. Effectiveness of homologous or heterologous immunization regimens against SARS-CoV-2 after two doses of inactivated COVID-19 vaccine: A systematic review and meta-analysis. Human Vaccines & Immunotherapeutics 19, 2221146. ISSN: 2164-5515. https://www.ncbi.nlm.nih.gov/pmc/articles/PMC10288895/ (2025) (June 2023).

67. Cromer, D. et al. Neutralising antibody titres as predictors of protection against SARS-CoV-2 variants and the impact of boosting: a meta-analysis. English. The Lancet Microbe 3. Publisher: Elsevier, e52–e61. ISSN: 2666-5247. https://www.thelancet.com/journals/lanmic/article/PIIS2666-5247(21)00267-6/fulltext (2025) (Jan. 2022).

68. Goel, R. R. et al. Efficient recall of Omicron-reactive B cell memory after a third dose of SARS-CoV-2 mRNA vaccine. Cell 185, 1875–1887.e8. ISSN: 0092-8674. https://www.sciencedirect.com/science/article/pii/S0092867422004561 (2025)(May 2022).

69. Rodda, L. B. et al. Imprinted SARS-CoV-2-specific memory lymphocytes define hybrid immunity. Cell 185, 1588–1601.e14. ISSN: 0092-8674. https://www.sciencedirect.com/science/article/pii/S0092867422003282 (2025) (Apr. 2022).

70. Blommaert, A. et al. The cost-effectiveness of pneumococcal vaccination in healthy adults over 50: An exploration of influential factors for Belgium. Vaccine 34, 2106–2112. ISSN: 0264-410X. https://www.sciencedirect.com/science/article/pii/S0264410×16002814 (2025) (Apr. 2016).

71. The World Health Organization. WHO SAGE Roadmap for prioritizing uses of COVID-19 vaccines: An approach to optimize the global impact of COVID-19 vaccines, based on public health goals, global and national equity, and vaccine access and coverage scenarios en. Oct. 2023. https://www.who.int/publications/i/item/WHO-2019-nCoV-Vaccines-SAGE-Prioritization-2023.1 (2025).

72. Wheatley, A. K. et al. Evolution of immune responses to SARS-CoV-2 in mild-moderate COVID-19. en. Nature Communications 12. Publisher: Nature Publishing Group, 1162. ISSN: 2041-1723. https://www.nature.com/articles/s41467-021-21444-5 (2025) (Feb. 2021).

73. Turner, J. S. et al. SARS-CoV-2 infection induces long-lived bone marrow plasma cells in humans. en. Nature 595. Publisher: Nature Publishing Group, 421–425. ISSN: 1476-4687. https://www.nature.com/articles/s41586-021-03647-4 (2025) (July 2021).

74. Mamais, I. et al. Circulating IgG Levels in SARS-CoV-2 Convalescent Individuals in Cyprus. en. Journal of Clinical Medicine 10. Number: 24 Publisher: Multidisciplinary Digital Publishing Institute, 5882. ISSN: 2077-0383. https://www.mdpi.com/2077-0383/10/24/5882 (2025) (Jan. 2021).

75. Akkaya, M., Kwak, K., & Pierce, S. K. B cell memory: building two walls of protection against pathogens. en. Nature Reviews Immunology 20. Number: 4 Publisher: Nature Publishing Group, 229–238. ISSN: 1474-1741. https://www.nature.com/articles/s41577-019-0244-2 (2023) (Apr. 2020).

76. Charles A Janeway, J., Travers, P., Walport, M., & Shlomchik, M. J. en. in Immunobiology: The Immune System in Health and Disease. 5th edition (Garland Science, 2001). https://www.ncbi.nlm.nih.gov/books/NBK27158/ (2024).

77. Tellier, J. & Nutt, S. L. Plasma cells: The programming of an antibody-secreting machine. en. European Journal of Immunology 49, 30–37. ISSN: 1521-4141. https://onlinelibrary.wiley.com/doi/abs/10.1002/eji.201847517 (2023) (2019).

78. Palm, A.-K. E. & Henry, C. Remembrance of Things Past: Long-Term B Cell Memory After Infection and Vaccination. Frontiers in Immunology 10. ISSN: 1664-3224. https://www.frontiersin.org/articles/10.3389/fimmu.2019.01787 (2023) (2019). 1

79. Administration (TGA), T. G. COVID-19 vaccines regulatory status | Therapeutic Goods Administration (TGA) en. text. Publisher: Therapeutic Goods Administration (TGA). June 2022. https://www.tga.gov.au/products/covid-19/covid-119-vaccines/covid-19-vaccines-regulatory-status (2025).

80. Trieu, M.-C. et al. Bivalent mRNA booster vaccination recalls cellular and antibody immunity against antigenically divergent SARS-CoV-2 spike antigens. en. npj Vaccines 10.Publisher: Nature Publishing Group, 74. ISSN: 2059-0105. https://www.nature.com/articles/s41541-025-01129-6 (2025) (Apr. 2025).

81. Voss, W. N. et al. Hybrid immunity to SARS-CoV-2 arises from serological recall of IgG antibodies distinctly imprinted by infection or vaccination. English. Cell Reports Medicine 5. Publisher: Elsevier. ISSN: 2666-3791. https://www.cell.com/cell-reports-medicine/abstract/S2666-3791(24)00382-3 (2025) (Aug. 2024).

82. Reynolds, C. J. et al. Heterologous infection and vaccination shapes immunity against SARS-CoV-2 variants. Science 375. Publisher: American Association for the Advancement of Science, 183–192. https://www.science.org/doi/full/10.1126/science.abm0811 (2025) (Jan. 2022).

83. Wheatley, A. K. et al. Immune imprinting and SARS-CoV-2 vaccine design. English. Trends in Immunology 42. Publisher: Elsevier, 956–959. ISSN: 1471-4906, 1471-4981. https://www.cell.com/trends/immunology/abstract/S1471-4906(21)00177-0 (2025) (Nov. 2021).

84. Huang, C. Q., Vishwanath, S., Carnell, G. W., Chan, A. C. Y., & Heeney, J. L. Immune imprinting and next-generation coronavirus vaccines. en. Nature Microbiology 8. Publisher: Nature Publishing Group, 1971–1985. ISSN: 2058-5276. https://www.nature.com/articles/s41564-023-01505-9 (2025) (Nov. 2023).

85. Koutsakos, M. & Ellebedy, A. H. Immunological imprinting: Understanding COVID-19. Immunity 56, 909–913. ISSN: 1074-7613. https://www.sciencedirect.com/science/article/pii/S1074761323001814 (2025) (May 2023).

86. Khoury, D. S. et al. Neutralizing antibody levels are highly predictive of immune protection from symptomatic SARS-CoV-2 infection. en. Nature Medicine 27. Publisher: Nature Publishing Group, 1205–1211. ISSN: 1546-170X. https://www.nature.com/articles/s41591-021-01377-8 (2024) (July 2021).

87. Goldblatt, D., Alter, G., Crotty, S., & Plotkin, S. A. Correlates of protection against SARS-CoV-2 infection and COVID-19 disease. en. Immunological Reviews 310. _eprint: https://onlinelibrary.wiley.com/doi/pdf/10.1111/imr.13091, 6–26. ISSN: 1600-065X. https://onlinelibrary.wiley.com/doi/abs/10.1111/imr.13091 (2025)(2022).

88. Khoury, D. S. et al. Correlates of Protection, Thresholds of Protection, and Immunobridging among Persons with SARS-CoV-2 Infection. Emerging Infectious Diseases 29, 381–388. ISSN: 1080-6040. https://www.ncbi.nlm.nih.gov/pmc/articles/PMC9881762/ (2025) (Feb. 2023).

89. Chen, W. et al. The kinetics of IgG subclasses and contributions to neutralizing activity against SARS-CoV-2 wild-type strain and variants in healthy adults immunized with inactivated vaccine. Immunology, 10.1111/imm.13531. ISSN: 0019-2805. https://www.ncbi.nlm.nih.gov/pmc/articles/PMC9349727/ (2025) (July 2022).

90. Dolscheid-Pommerich, R. et al. Correlation between a quantitative anti-SARS-CoV-2 IgG ELISA and neutralization activity. en. Journal of Medical Virology 94. _eprint: https://onlinelibrary.wiley.com/doi/pdf/10.1002/jmv.27287, 388–392. ISSN: 1096-9071 https://onlinelibrary.wiley.com/doi/abs/10.1002/jmv.27287 (2025) (2022).

91. Higashimoto, Y. et al. Correlation between anti-S IgG and neutralizing antibody titers against three live SARS-CoV-2 variants in BNT162b2 vaccine recipients. Human Vaccines & Immunotherapeutics 18. Publisher: Taylor & Francis, 2105611. ISSN: 2164-5515. 10.1080/21645515.2022.2105611 (2025) (Nov. 2022).

92. Ramos, A., Cardoso, M. J., Ribeiro, L., & Guimarães, J. T. Assessing SARS-CoV-2 Neutralizing Antibodies after BNT162b2 Vaccination and Their Correlation with SARS-CoV-2 IgG Anti-S1, Anti-RBD and Anti-S2 Serological Titers. en. Diagnostics 12. Number: 1 Publisher: Multidisciplinary Digital Publishing Institute, 205. ISSN: 2075-4418. https://www.mdpi.com/2075-4418/12/1/205 (2025) (Jan. 2022).

93. Huang, A. T. et al. A systematic review of antibody mediated immunity to coronaviruses: kinetics, correlates of protection, and association with severity. en. Nature Communications 11. Publisher: Nature Publishing Group, 4704. ISSN: 2041-1723. https://www.nature.com/articles/s41467-020-18450-4 (2025) (Sept. 2020).

94. Goldblatt, D. et al. Towards a population-based threshold of protection for COVID-19 vaccines. Vaccine 40, 306–315. ISSN: 0264-410X. https://www.sciencedirect.com/science/article/pii/S0264410×21016157 (2025) (Jan. 2022).

95. Hay, J. A., Laurie, K., White, M., & Riley, S. Characterising antibody kinetics from multiple influenza infection and vaccination events in ferrets. en. PLOS Computational Biology 15. Publisher: Public Library of Science, e1007294. ISSN: 1553-7358. https://journals.plos.org/ploscompbiol/article?id=10.1371/journal.pcbi.1007294 (2023) (Aug. 2019).

96. Yan, A. W. C. et al. Modelling cross-reactivity and memory in the cellular adaptive immune response to influenza infection in the host. Journal of Theoretical Biology 413, 34–49. ISSN: 0022-5193. https://www.sciencedirect.com/science/article/pii/S0022519316303629 (2025) (Jan. 2017).

97. De Graaf, W. F., Kretzschmar, M. E. E., Teunis, P. F. M., & Diekmann, O. A two-phase within-host model for immune response and its application to serological profiles of pertussis. Epidemics 9, 1–7. ISSN: 1755-4365. https://www.sciencedirect.com/science/article/pii/S1755436514000371 (2025) (Dec. 2014).

