## Supplementary material for "Quantification of the IgG antibody response half-life for hybrid immunity to SARS-CoV-2"

### A. Supplementary material

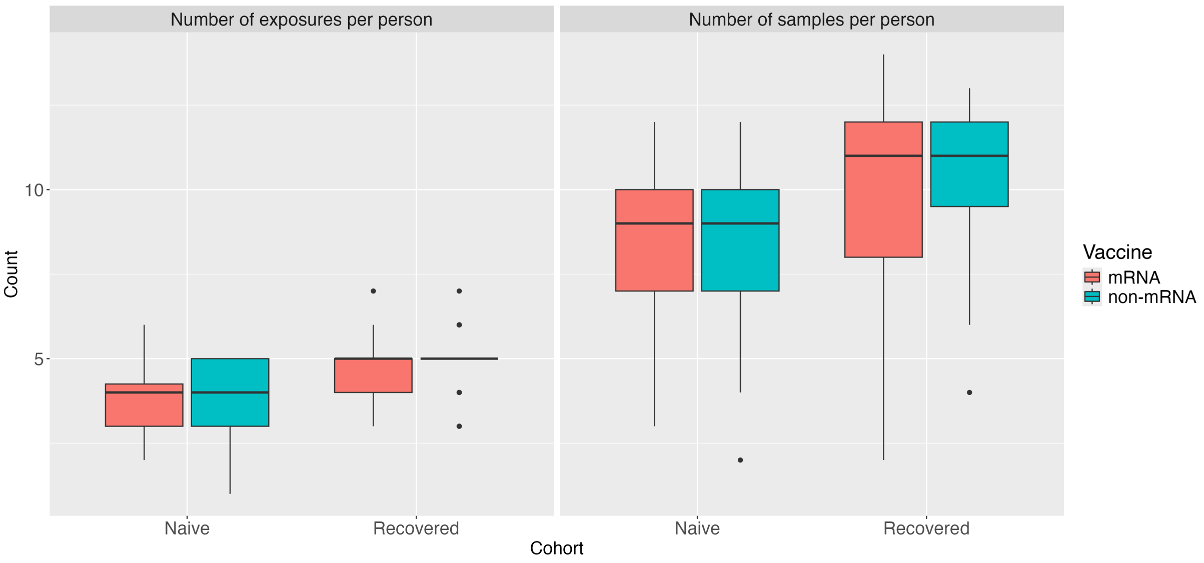


**Figure S1: The number of exposures experienced and samples collected across individuals in the naive and recovered cohorts.**

**Table S1: P-values from a t-test measuring the difference in geometric mean between the naive and recovered cohorts.**

| | Vaccine | First vaccination | Second vaccination | Booster vaccination |
|---|---|---|---|---|
| **Spike antigen** | mRNA | 0.0002797899 | 0.0017343010 | 0.1828342621 |
| | non-mRNA | 0.0002929919 | 0.0007801639 | 0.3375290516 |
| **RBD antigen** | mRNA | 0.0001690447 | 0.1010475029 | 0.2308697530 |
| | non-mRNA | 0.0017179875 | 0.0012199818 | 0.7435133242 |

**Table S2: P-values from a t-test measuring the difference in geometric mean between the mRNA and non-mRNA recipients.**

| | Cohort | First vaccination | Second vaccination | Booster vaccination |
|---|---|---|---|---|
| **Spike antigen** | Naive | 0.078905759 | 0.003780063 | 0.250280466 |
| | Recovered | 0.114733486 | 0.008450363 | 0.503777106 |
| **RBD antigen** | Naive | 0.84441012 | 0.47303610 | 0.23492083 |
| | Recovered | 0.20990570 | 0.03788605 | 0.78502580 |

**Table S3: P-values from a t-test measuring the difference in geometric mean between vaccination events.**

| | Cohort | mRNA | | non-mRNA | |
|---|---|---|---|---|---|
| | | V1-V2 | V2-B1 | V1-V2 | V2-B1 |
| **Spike antigen** | Naive | 0.004049353 | 0.7770513821 | 0.005432911 | 0.0002039171 |
| | Recovered | 0.509509334 | 0.0100584512 | 0.663147098 | 0.0620381004 |
| **RBD antigen** | Naive | 0.002516619 | 0.9978369904 | 0.003233262 | 0.0002600634 |
| | Recovered | 0.625644766 | 0.3107499487 | 0.429290729 | 0.1126627850 |

#### Table S4: Posterior population-level parameter median estimates and 95% credible intervals for the IgG response to spike

| | Vaccine type | Population-level parameter estimates to spike | | Population-level parameter estimates to RBD | |
|---|---|---|---|---|---|
| | | $\mu_{d_\lambda}$ | $\phi$ | $\mu_{d_\lambda}$ | $\phi$ |
| **Naive cohort** | mRNA | 220.102<br>[178.684, 281.89] | 0.003<br>[0.003, 0.004] | 177.877<br>[146.464, 227.846] | 0.003<br>[0.002, 0.004] |
| | non-mRNA | 210.665<br>[166.361, 282.563] | 0.004<br>[0.003, 0.005] | 201.896<br>[157.897, 275.436] | 0.003<br>[0.003, 0.005] |
| **Recovered cohort** | mRNA | 452.55<br>[376.377, 556.345] | 0.006<br>[0.005, 0.007] | 499.045<br>[404.911, 635.787] | 0.006<br>[0.005, 0.007] |
| | non-mRNA | 488.39<br>[372.377, 655.727] | 0.007<br>[0.005, 0.008] | 452.228<br>[344.476, 609.822] | 0.008<br>[0.006, 0.01] |
| | None | 318.094<br>[269.609, 386.806] | 0.021<br>[0.017, 0.026] | 343.383<br>[285.434, 427.258] | 0.025<br>[0.02, 0.031] |

#### Table S5: Posterior population-level parameter median estimates and 95% credible intervals for the IgG response to spike

| | Vaccine type | Population-level parameter estimates | | | | | | |
|---|---|---|---|---|---|---|---|---|
| | | $\mu_\beta$ | $\mu_\rho$ | $\mu_{d_a}$ | $\mu_{d_s}$ | $\mu_{d_l}$ | $\mu_m$ | $\phi$ |
| **Naive cohort** | mRNA | 1732.139<br>[1360.321, 2265.412] | 0.882<br>[0.833, 0.917] | 21.162<br>[17.394, 25.677] | 4.735<br>[3.613, 6.2] | 450.587<br>[336.033, 620.008] | 0.509<br>[0.368, 0.664] | 0.003<br>[0.002, 0.004] |
| | non-mRNA | 1012.643<br>[624.495, 1736.917] | 0.835<br>[0.596, 0.899] | 19.774<br>[16.179, 24.175] | 3.543<br>[2.652, 4.758] | 395.19<br>[282.631, 566.085] | 1.469<br>[1.088, 1.865] | 0.003<br>[0.002, 0.005] |
| **Recovered cohort** | mRNA | 2266.505<br>[1701.582, 3063.617] | 0.886<br>[0.842, 0.917] | 19.486<br>[16.089, 23.656] | 3.55<br>[2.806, 4.503] | 859.19<br>[685.72, 1095.444] | 0.351<br>[0.22, 0.516] | 0.006<br>[0.005, 0.007] |
| | non-mRNA | 2011.675<br>[1416.902, 2935.789] | 0.865<br>[0.8, 0.906] | 20.557<br>[16.88, 24.931] | 4.176<br>[3.175, 5.495] | 852.522<br>[629.357, 1138.371] | 0.445<br>[0.268, 0.687] | 0.005<br>[0.004, 0.007] |
| | None | 1383.015<br>[843.777, 2281.168] | 0.754<br>[0.569, 0.865] | 19.236<br>[15.99, 23.272] | 3.665<br>[2.733, 4.918] | 495.243<br>[380.687, 661.212] | | 0.008<br>[0.006, 0.011] |

#### Table S6: Posterior population-level parameter median estimates and 95% credible intervals for the IgG response to RBD

| | Vaccine type | Population-level parameter estimates | | | | | | |
|---|---|---|---|---|---|---|---|---|
| | | $\mu_\beta$ | $\mu_\rho$ | $\mu_{d_a}$ | $\mu_{d_s}$ | $\mu_{d_l}$ | $\mu_m$ | $\phi$ |
| **Naive cohort** | mRNA | 1007.05<br>[708.697, 1540.897] | 0.885<br>[0.816, 0.924] | 20.754<br>[16.943, 25.343] | 4.339<br>[3.221, 5.82] | 431.709<br>[304.191, 614.215] | 1.311<br>[1.089, 1.54] | 0.002<br>[0.001, 0.003] |
| | non-mRNA | 1060.181<br>[638.282, 1854.156] | 0.778<br>[0.505, 0.881] | 19.78<br>[16.208, 24.108] | 3.385<br>[2.544, 4.526] | 375.853<br>[268.801, 539.362] | 1.57<br>[1.113, 2.063] | 0.004<br>[0.003, 0.006] |
| **Recovered cohort** | mRNA | 1898.263<br>[1421.44, 2605.016] | 0.834<br>[0.748, 0.885] | 18.091<br>[14.913, 21.968] | 3.409<br>[2.654, 4.367] | 807.758<br>[645.398, 1030.91] | 0.351<br>[0.219, 0.519] | 0.005<br>[0.004, 0.006] |
| | non-mRNA | 1743.041<br>[1211.244, 2581.651] | 0.841<br>[0.723, 0.895] | 20.725<br>[16.762, 25.576] | 4.035<br>[3.023, 5.415] | 773.317<br>[565.049, 1053.024] | 0.452<br>[0.269, 0.705] | 0.005<br>[0.004, 0.007] |
| | None | 1370.721<br>[854.379, 2222.954] | 0.753<br>[0.572, 0.863] | 19.367<br>[16.042, 23.486] | 3.64<br>[2.722, 4.888] | 435.88<br>[336.594, 586.509] | | 0.009<br>[0.007, 0.011] |

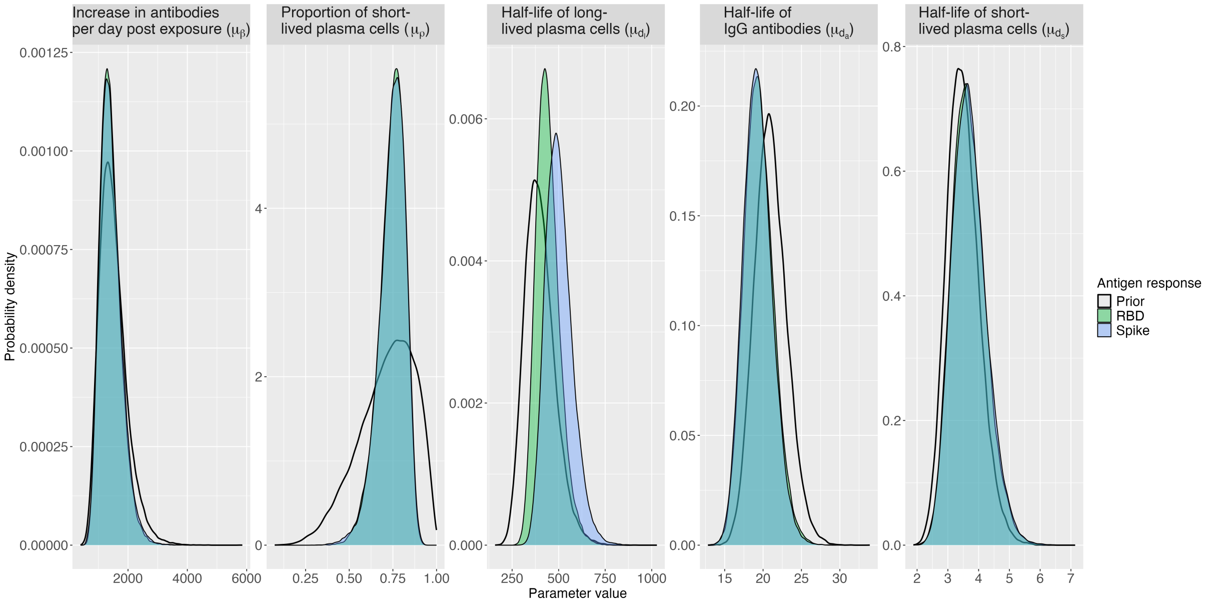


**Figure S2: Population-level parameter estimate distributions for the IgG response to RBD and spike protein.**

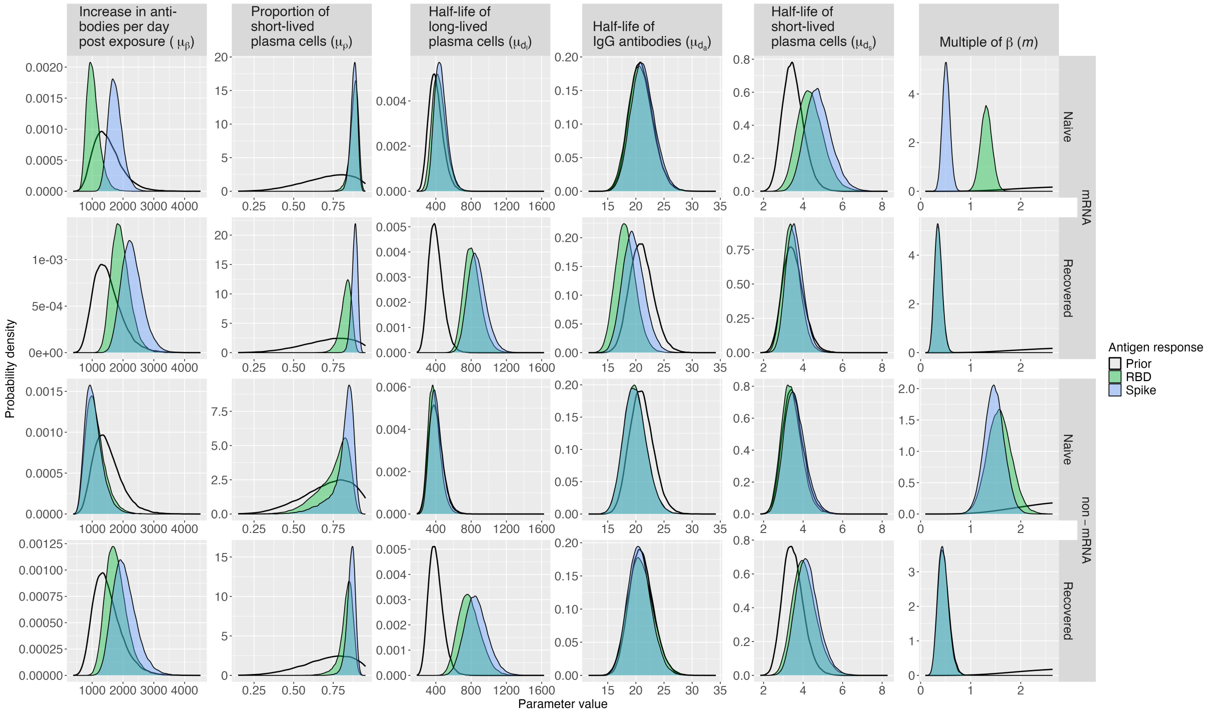


**Figure S3: Population-level parameter estimate distributions for the IgG response to RBD and spike protein.**

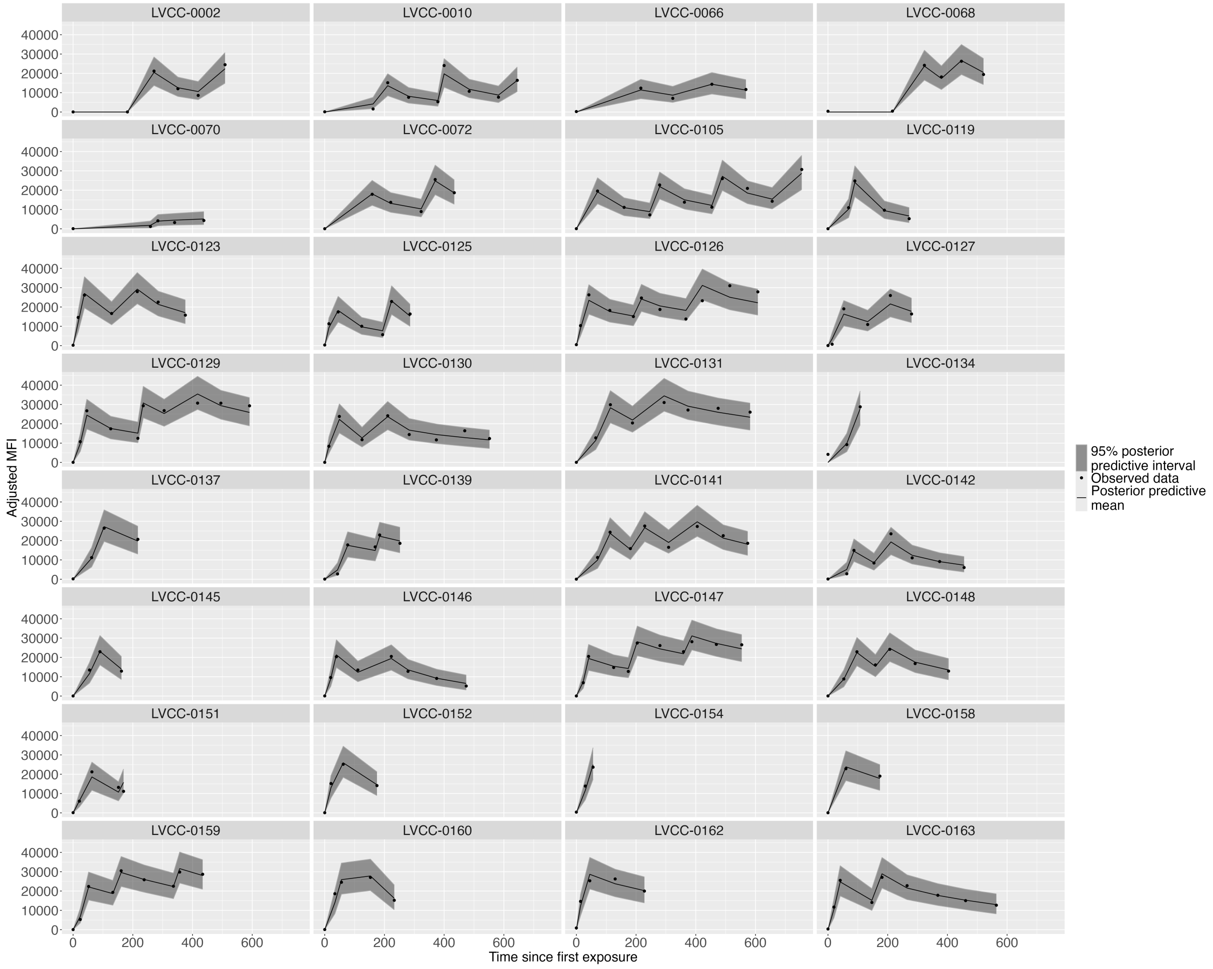


**Figure S4: Bi-phasic decay model fits with observed data for the IgG response to spike for naive mRNA recipients.** Posterior predictive means are represented by the dashed black line and the 95% posterior predictive interval is shown by the grey ribbon. Observed data is indicated by black dots.

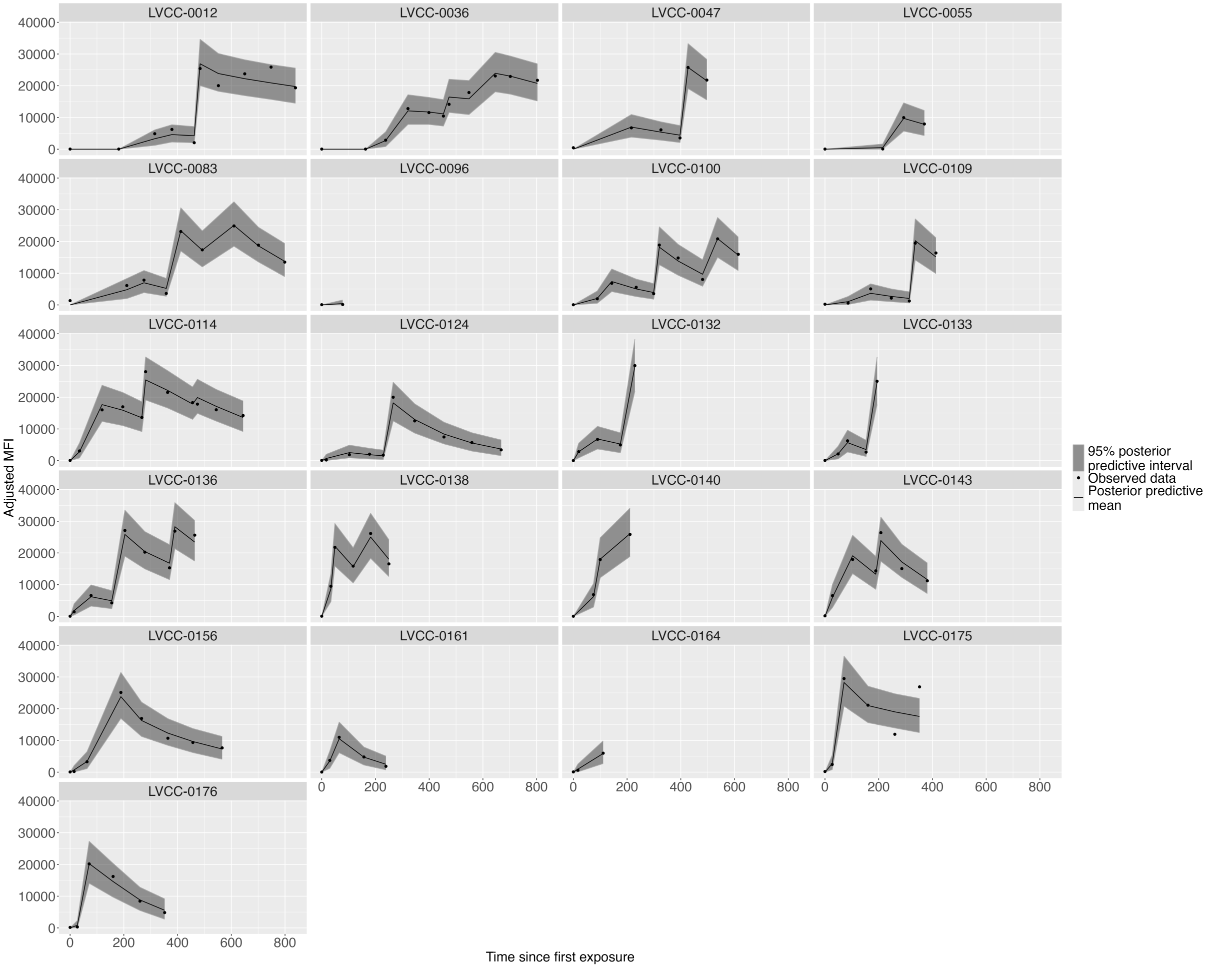


**Figure S5: Bi-phasic decay model fits with observed data for the IgG response to spike for naive non-mRNA recipients.** Posterior predictive means are represented by the dashed black line and the 95% posterior predictive interval is shown by the grey ribbon. Observed data is indicated by black dots.

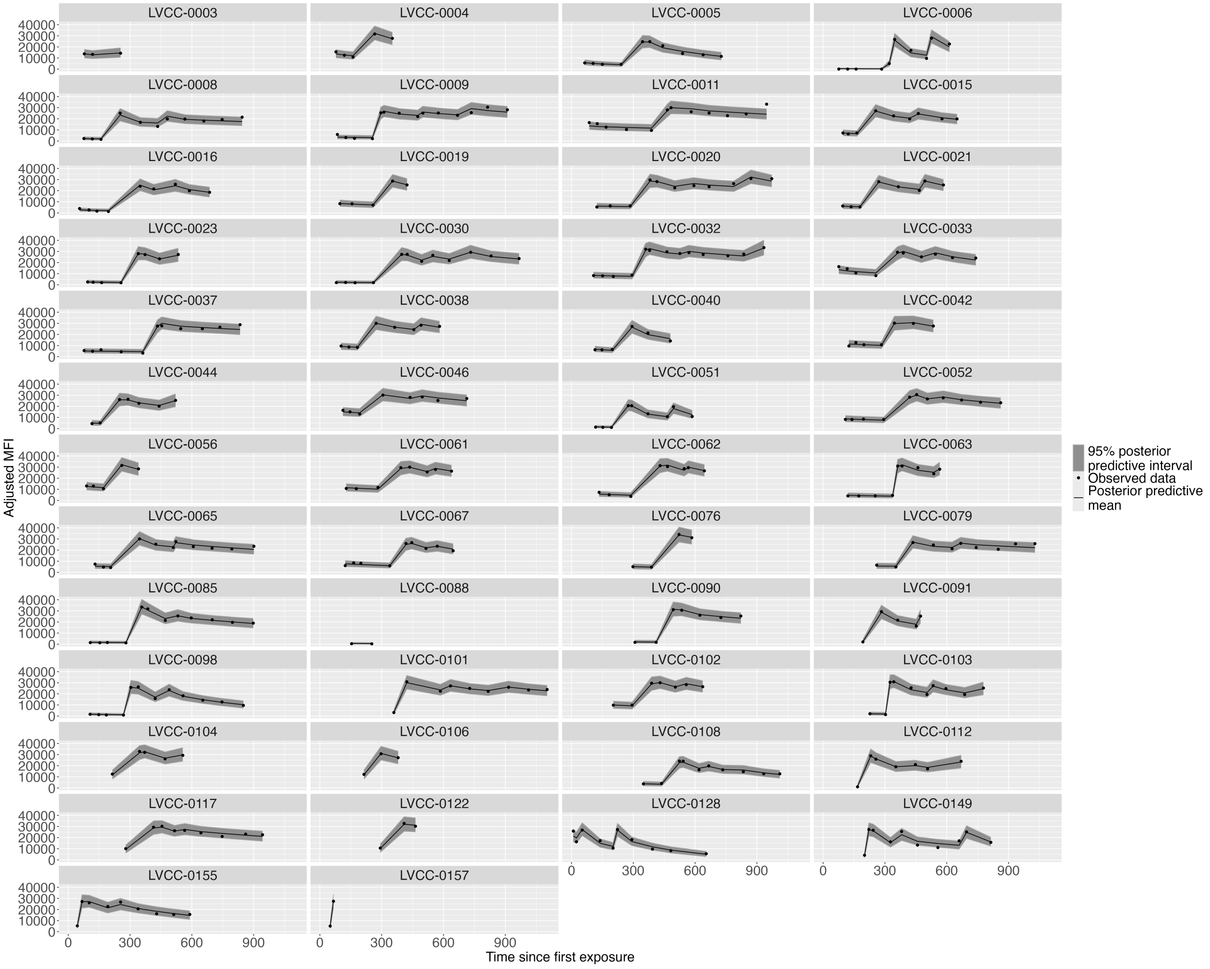


**Figure S6: Bi-phasic decay model fits with observed data for the IgG response to spike for recovered mRNA recipients.** Posterior predictive means are represented by the dashed black line and the 95% posterior predictive interval is shown by the grey ribbon. Observed data is indicated by black dots.

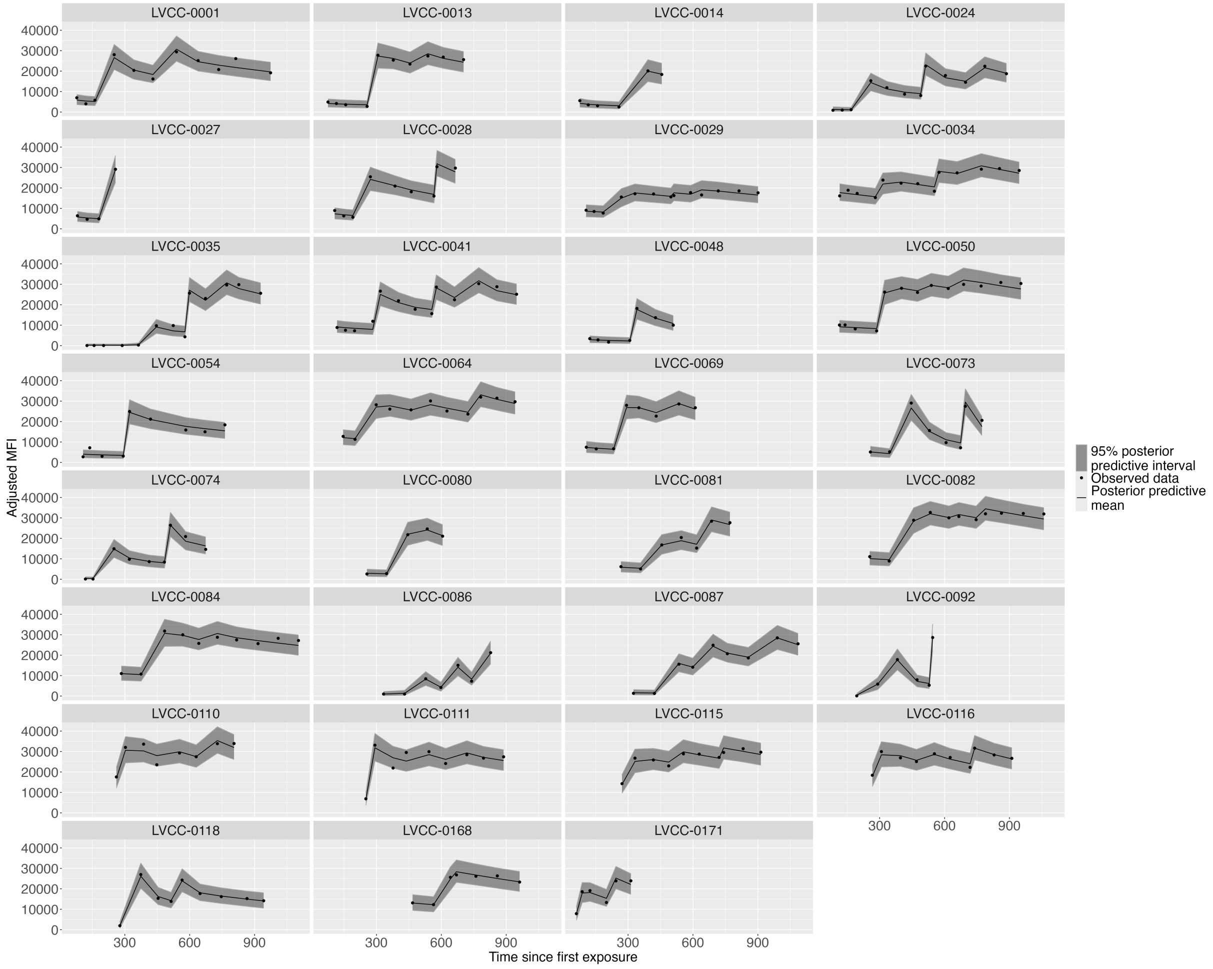


**Figure S7: Bi-phasic decay model fits with observed data for the IgG response to spike for recovered non-mRNA recipients.** Posterior predictive means are represented by the dashed black line and the 95% posterior predictive interval is shown by the grey ribbon. Observed data is indicated by black dots.

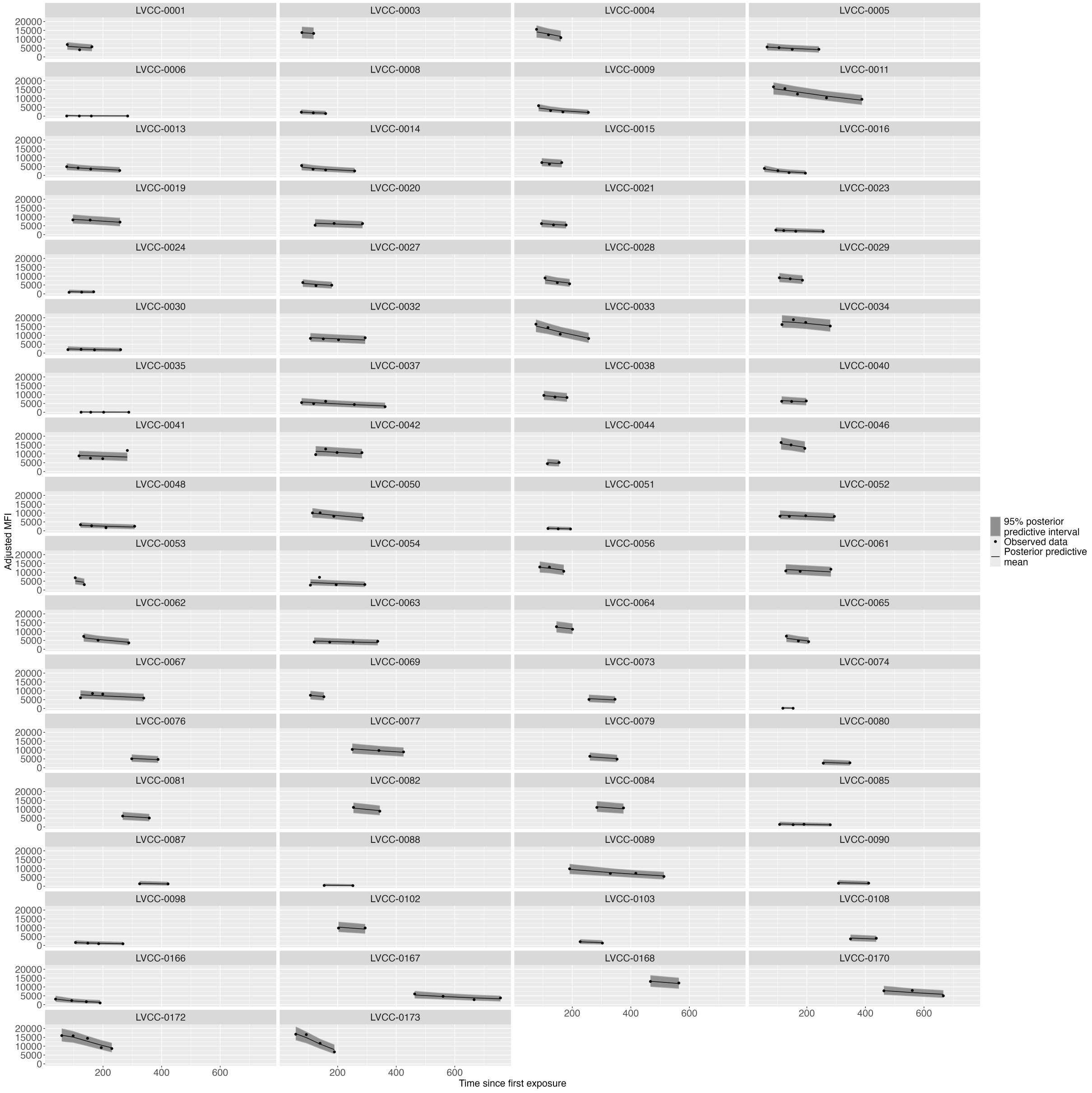


**Figure S8: Bi-phasic decay model fits with observed data for the IgG response to spike for recovered individuals before vaccination.** Posterior predictive means are represented by the dashed black line and the 95% posterior predictive interval is shown by the grey ribbon. Observed data is indicated by black dots.

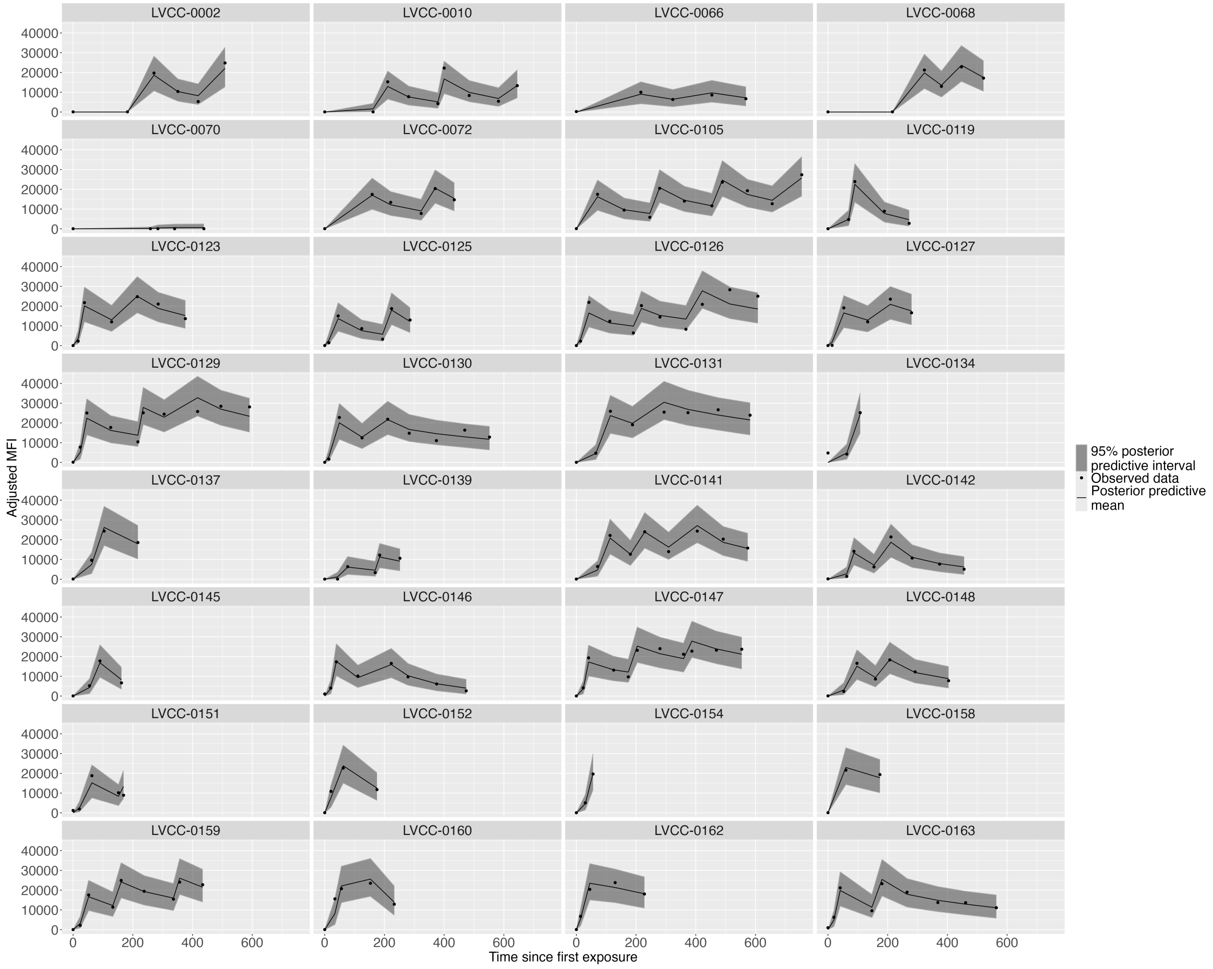


**Figure S9: Bi-phasic decay model fits with observed data for the IgG response to RBD for naive mRNA recipients.** Posterior predictive means are represented by the dashed black line and the 95% posterior predictive interval is shown by the grey ribbon. Observed data is indicated by black dots.

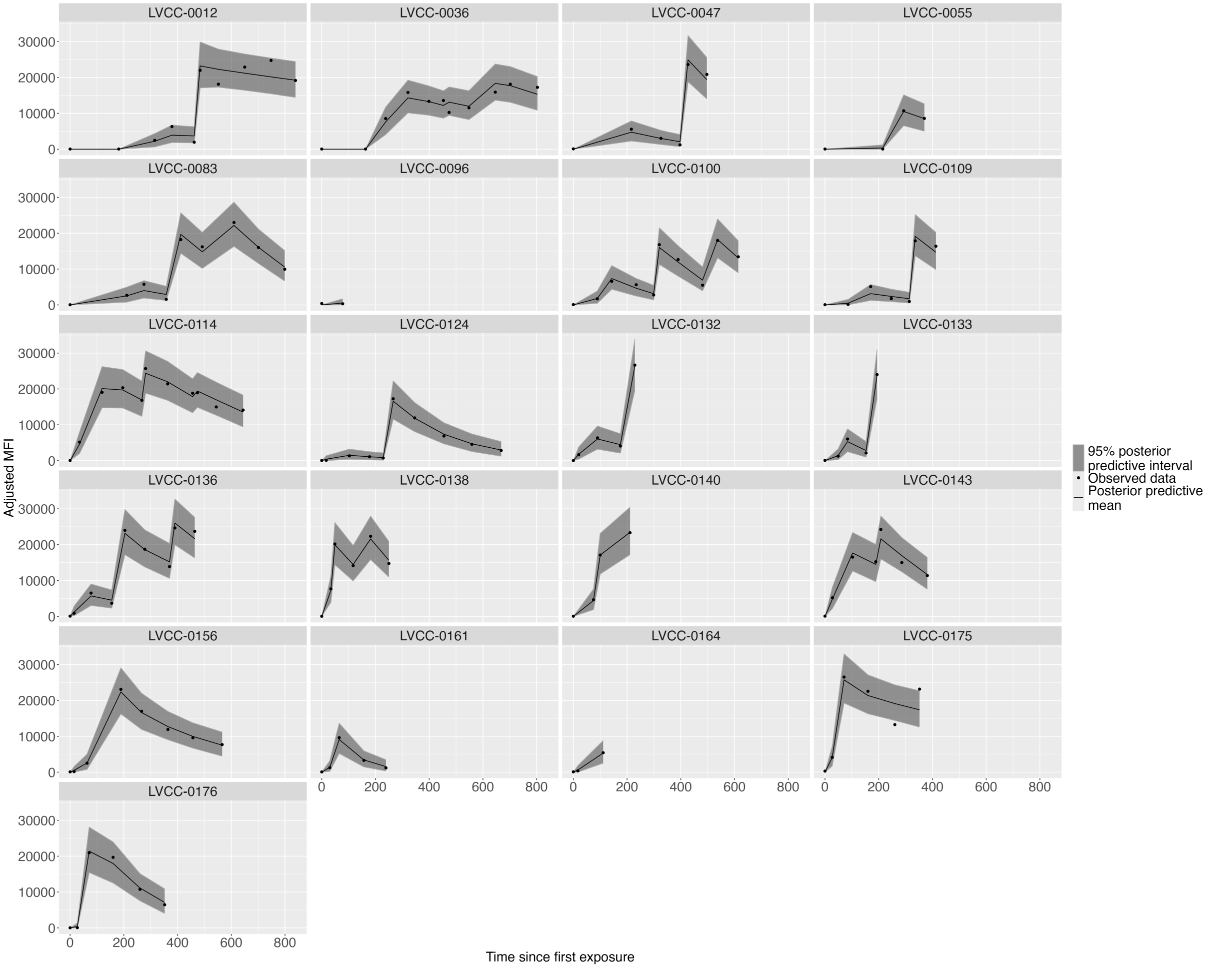


**Figure S10: Bi-phasic decay model fits with observed data for the IgG response to RBD for naive non-mRNA recipients.** Posterior predictive means are represented by the dashed black line and the 95% posterior predictive interval is shown by the grey ribbon. Observed data is indicated by black dots.

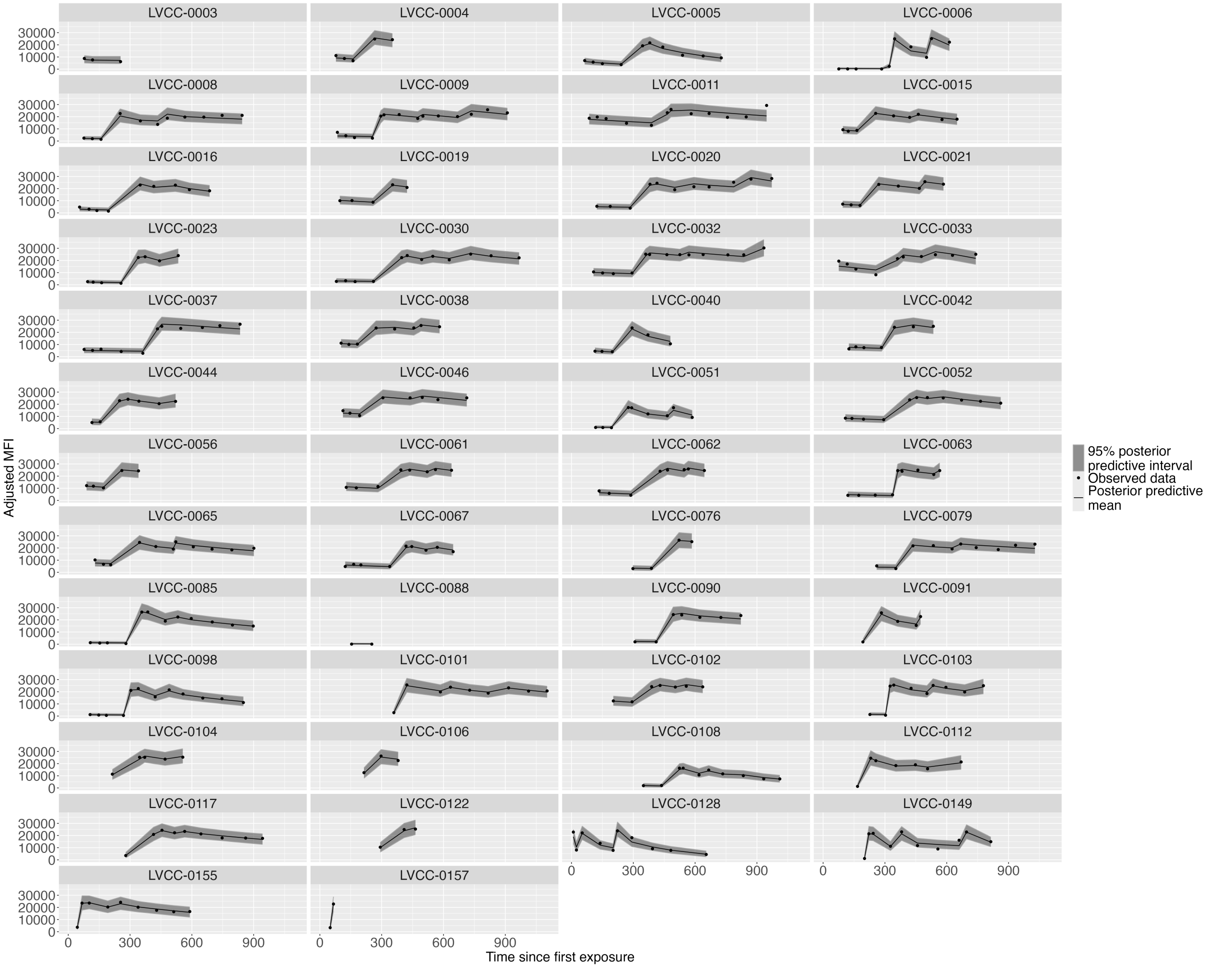


**Figure S11: Bi-phasic decay model fits with observed data for the IgG response to RBD for recovered mRNA recipients.** Posterior predictive means are represented by the dashed black line and the 95% posterior predictive interval is shown by the grey ribbon. Observed data is indicated by black dots.

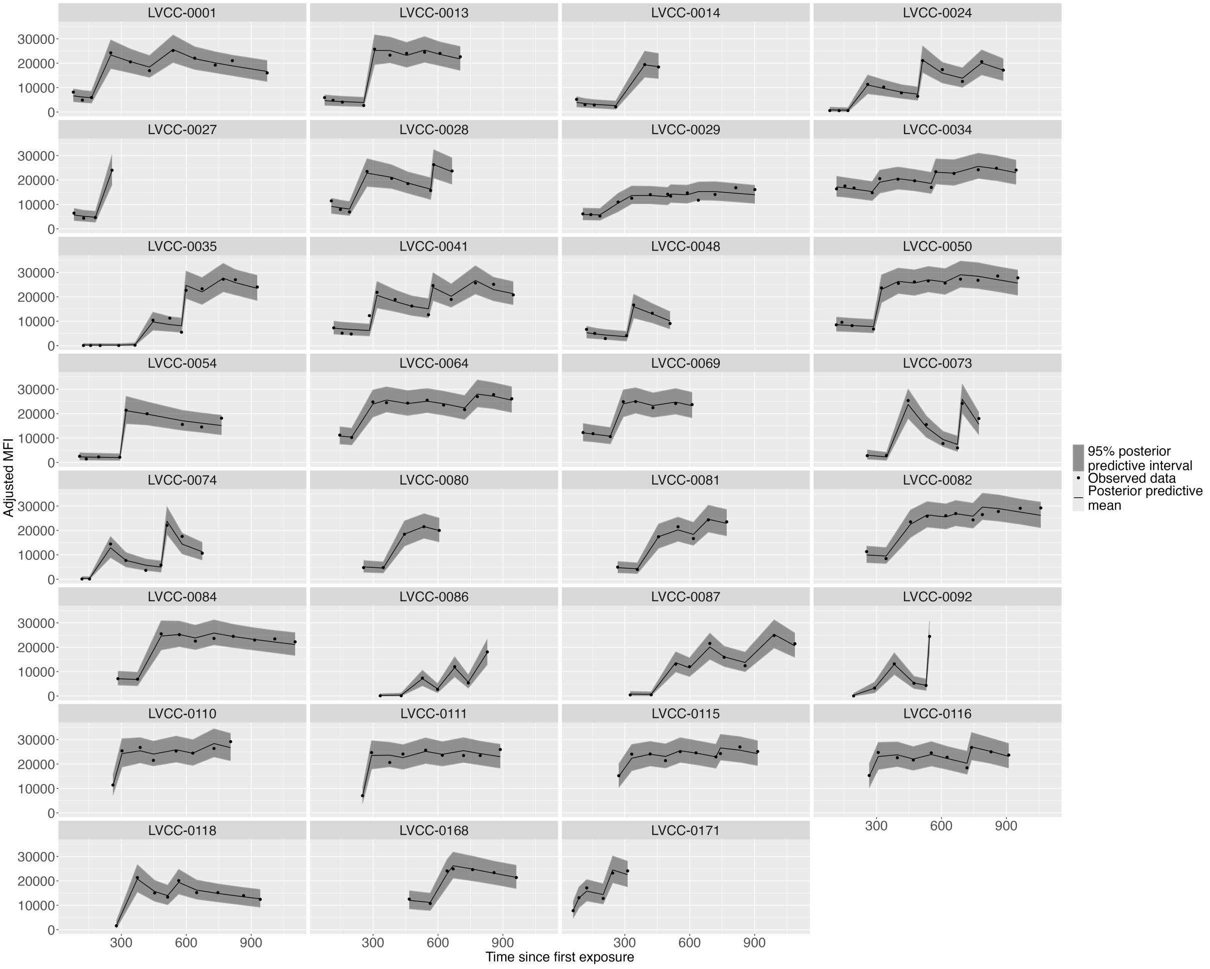


**Figure S12: Bi-phasic decay model fits with observed data for the IgG response to RBD for recovered non-mRNA recipients.** Posterior predictive means are represented by the dashed black line and the 95% posterior predictive interval is shown by the grey ribbon. Observed data is indicated by black dots.

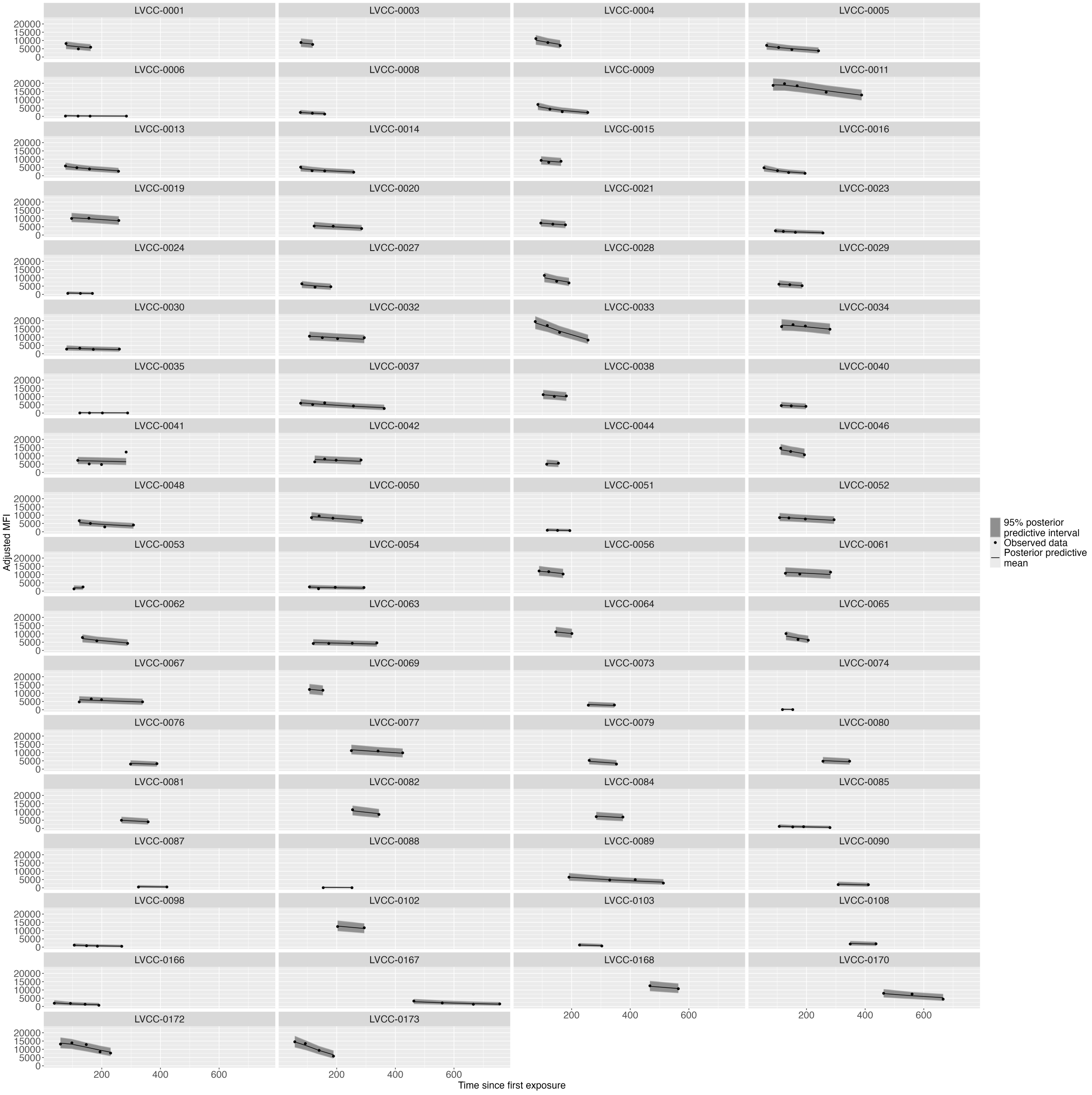


**Figure S13: Bi-phasic decay model fits with observed data for the IgG response to RBD for recovered individuals before vaccination.** Posterior predictive means are represented by the dashed black line and the 95% posterior predictive interval is shown by the grey ribbon. Observed data is indicated by black dots.

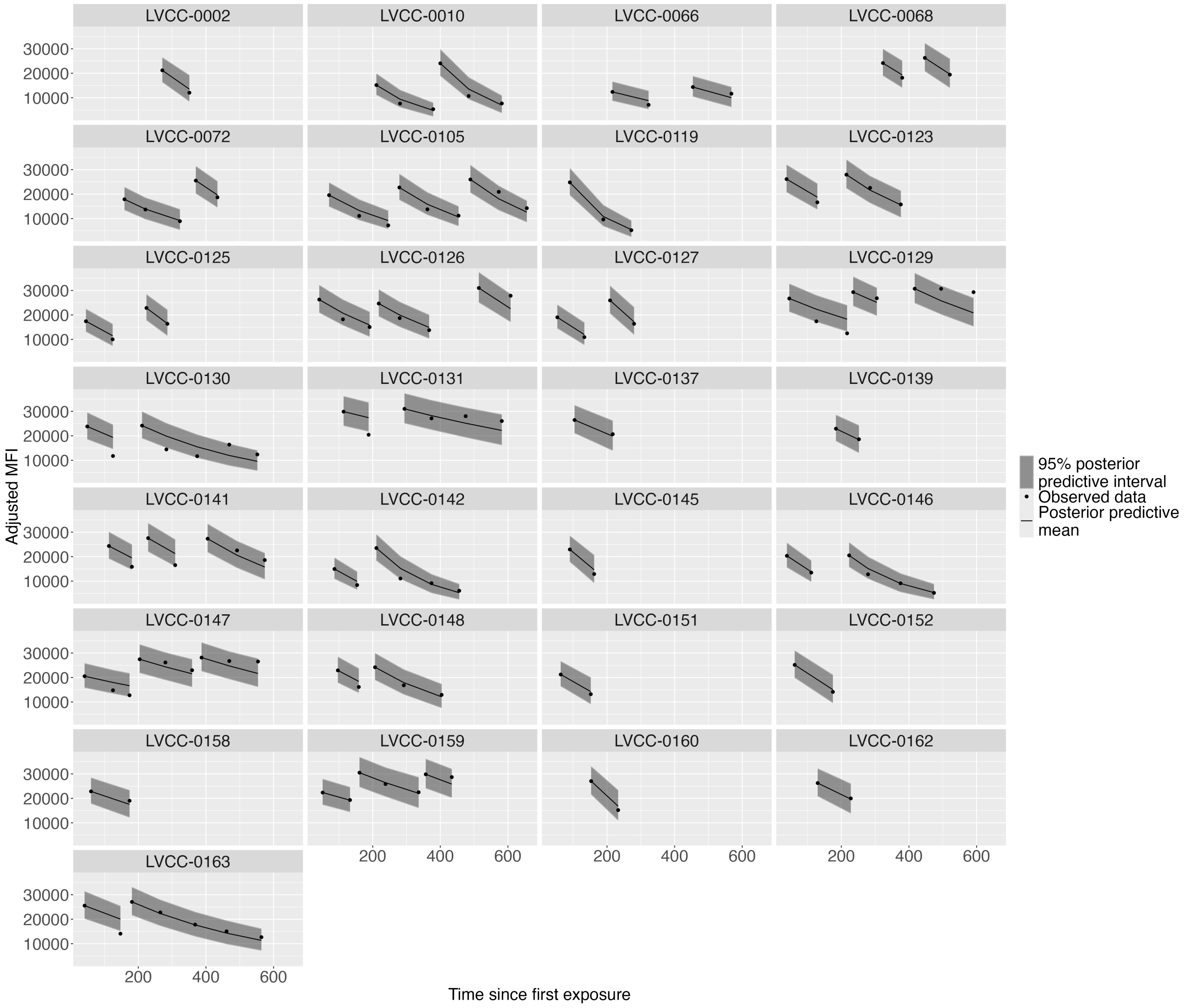


**Figure S14: Single phase decay model fits with observed data for the IgG response to spike for naive mRNA recipients.** Posterior predictive means are represented by the dashed black line and the 95% posterior predictive interval is shown by the grey ribbon. Observed data is indicated by black dots.

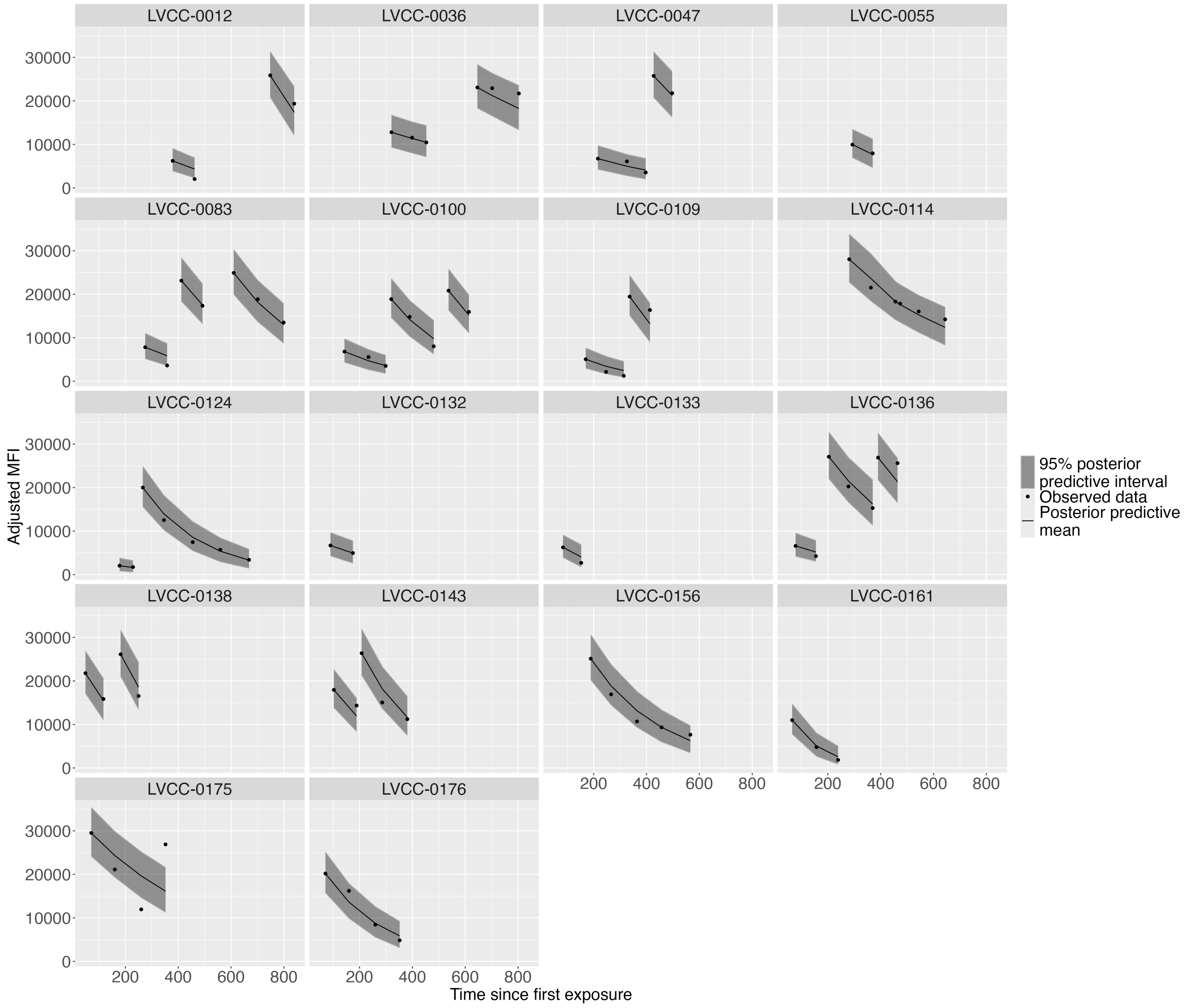


**Figure S15: Single phase decay model fits with observed data for the IgG response to spike for naive non-mRNA recipients.** Posterior predictive means are represented by the dashed black line and the 95% posterior predictive interval is shown by the grey ribbon. Observed data is indicated by black dots.

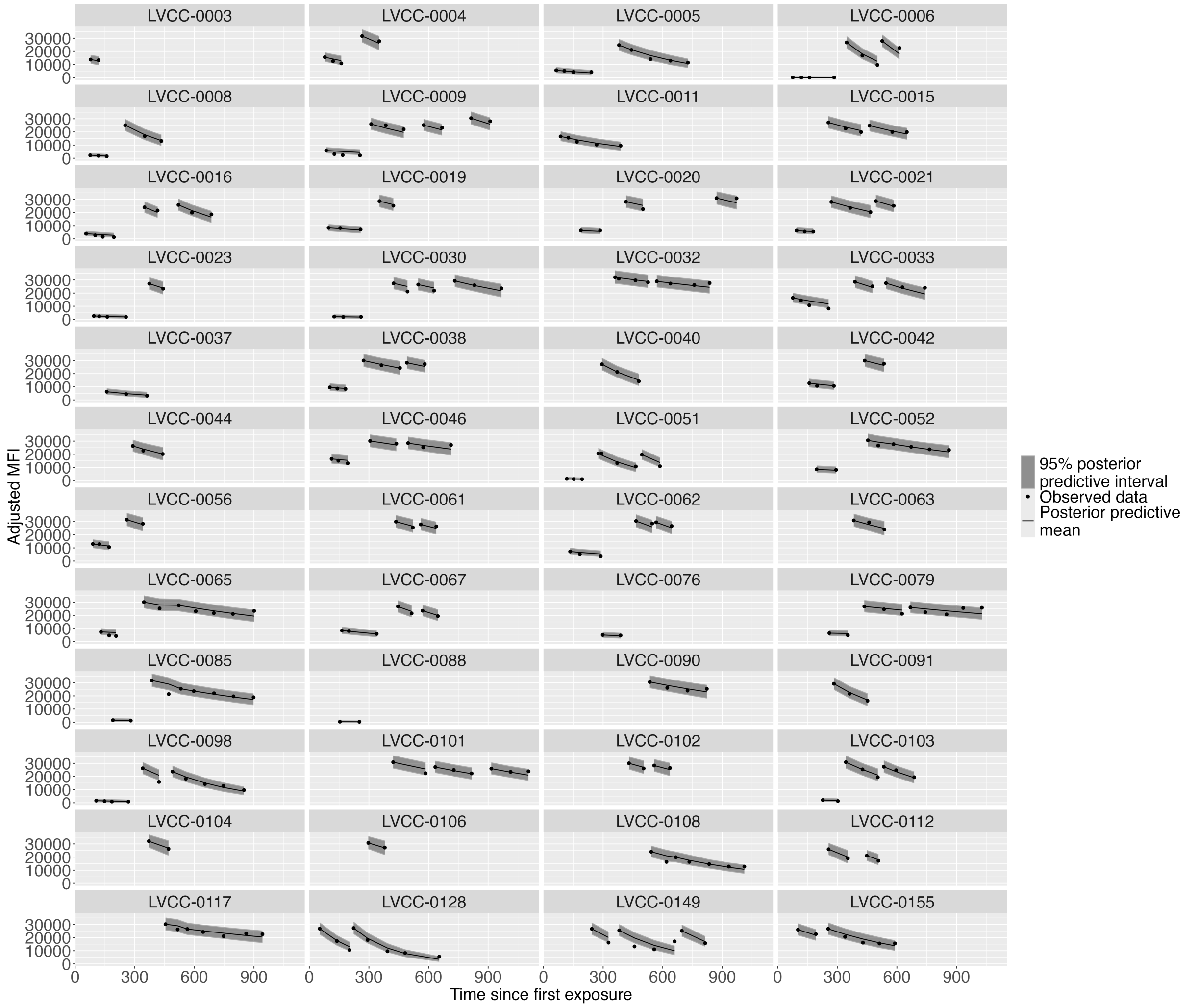


**Figure S16: Single phase decay model fits with observed data for the IgG response to spike for recovered mRNA recipients.** Posterior predictive means are represented by the dashed black line and the 95% posterior predictive interval is shown by the grey ribbon. Observed data is indicated by black dots.

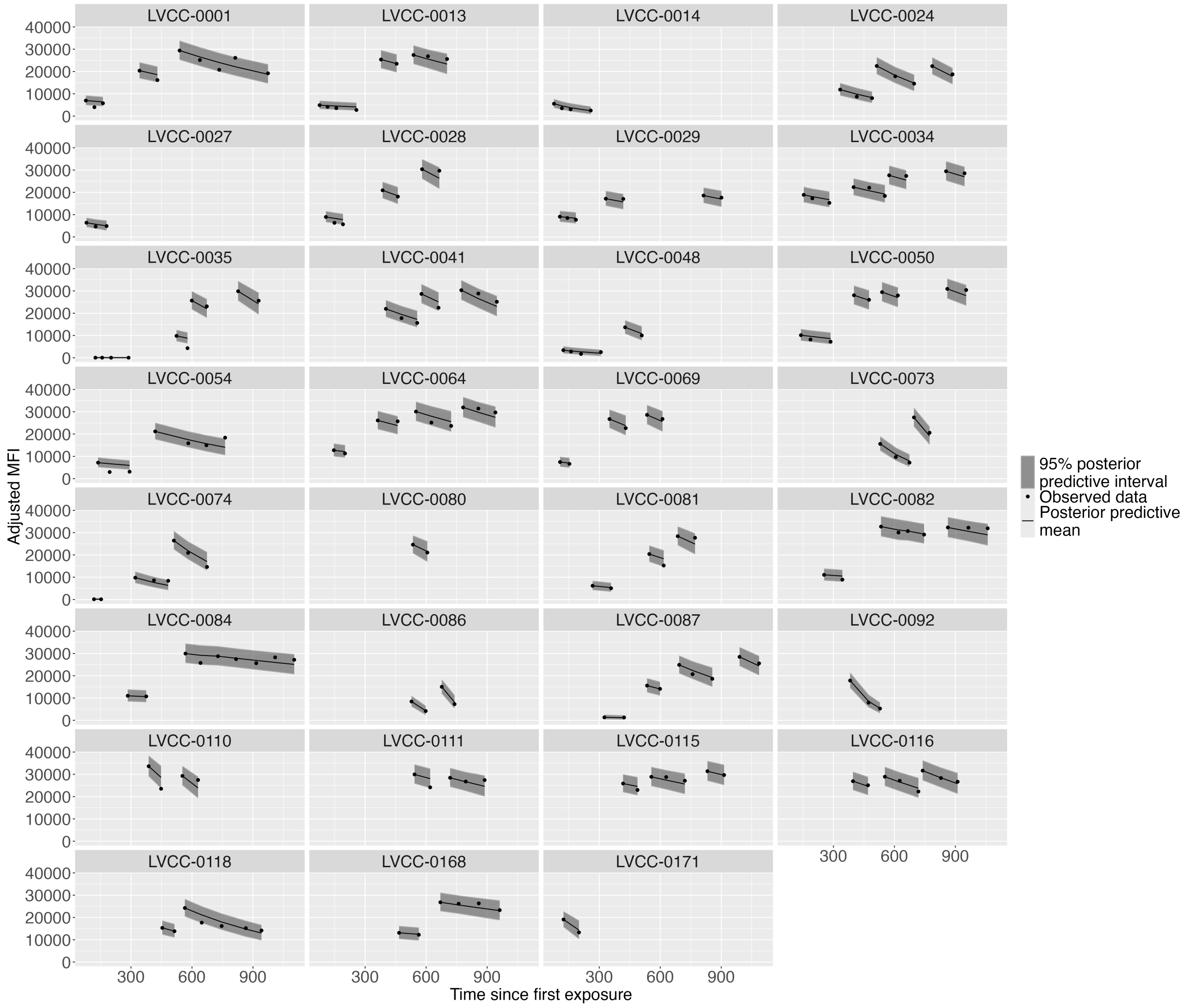


**Figure S17: Single phase decay model fits with observed data for the IgG response to spike for recovered non-mRNA recipients.** Posterior predictive means are represented by the dashed black line and the 95% posterior predictive interval is shown by the grey ribbon. Observed data is indicated by black dots.

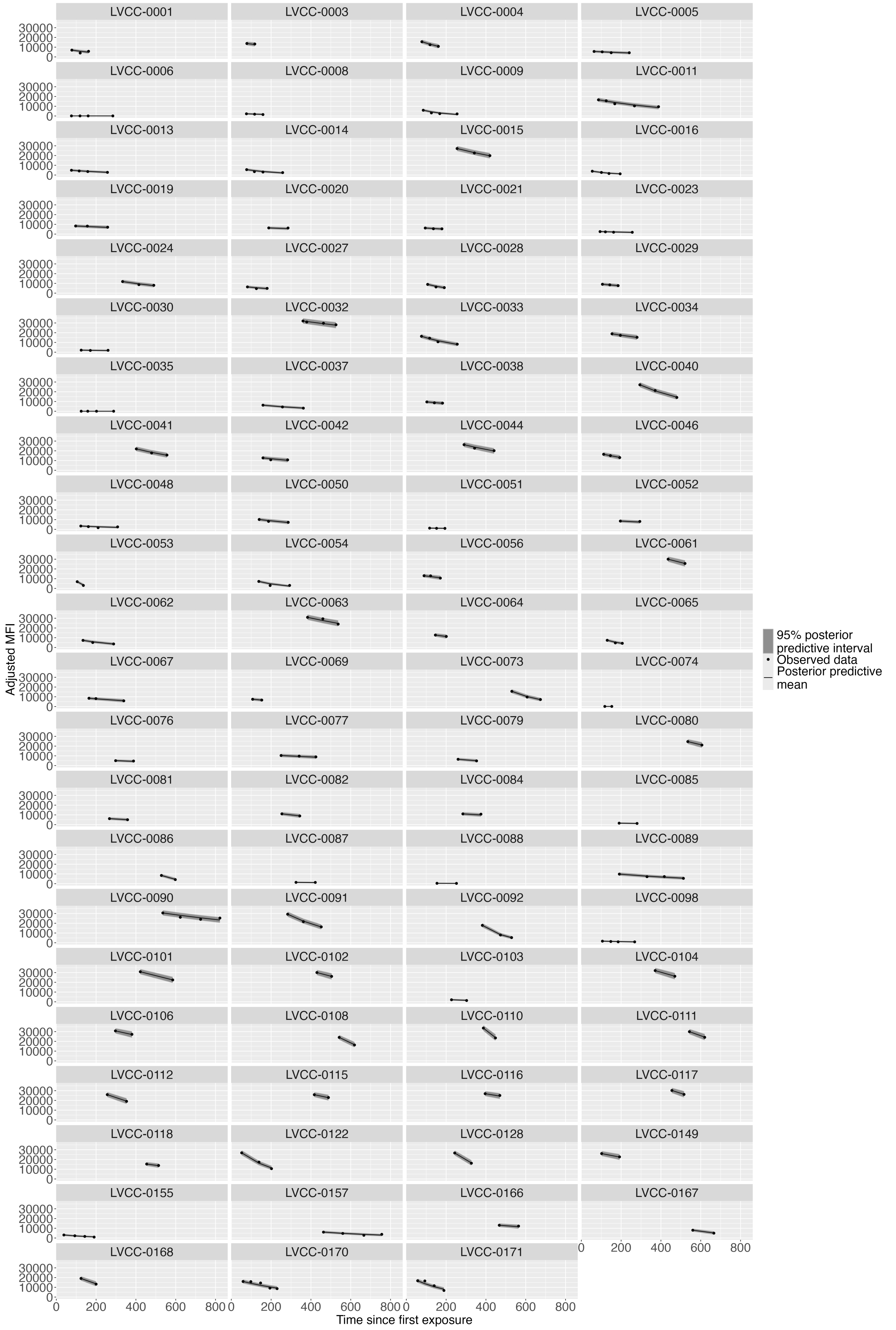


**Figure S18: Single phase decay model fits with observed data for the IgG response to spike for recovered individuals before vaccination.** Posterior predictive means are represented by the dashed black line and the 95% posterior predictive interval is shown by the grey ribbon. Observed data is indicated by black dots.

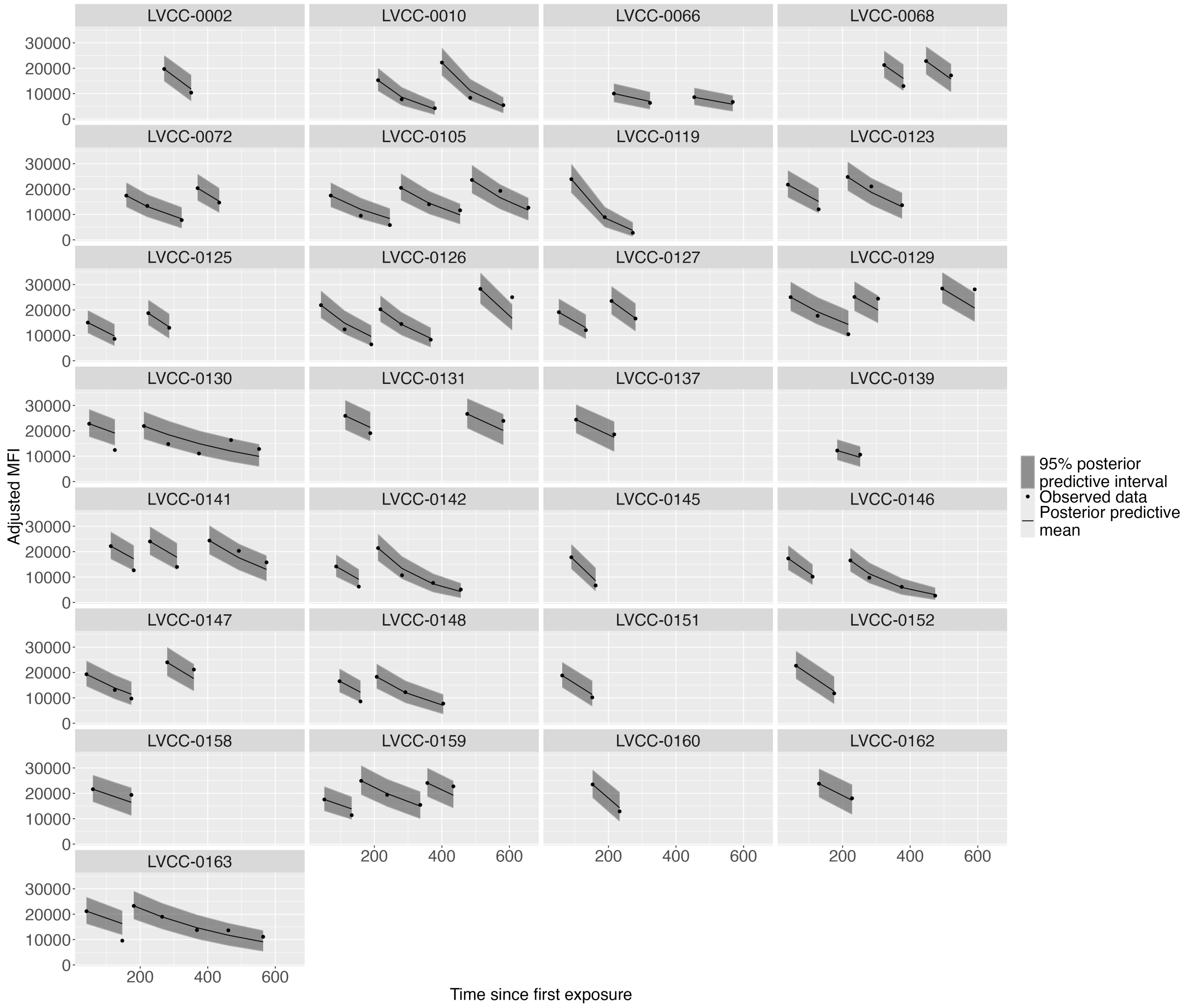


**Figure S19: Single phase decay model fits with observed data for the IgG response to RBD for naive mRNA recipients.** Posterior predictive means are represented by the dashed black line and the 95% posterior predictive interval is shown by the grey ribbon. Observed data is indicated by black dots.

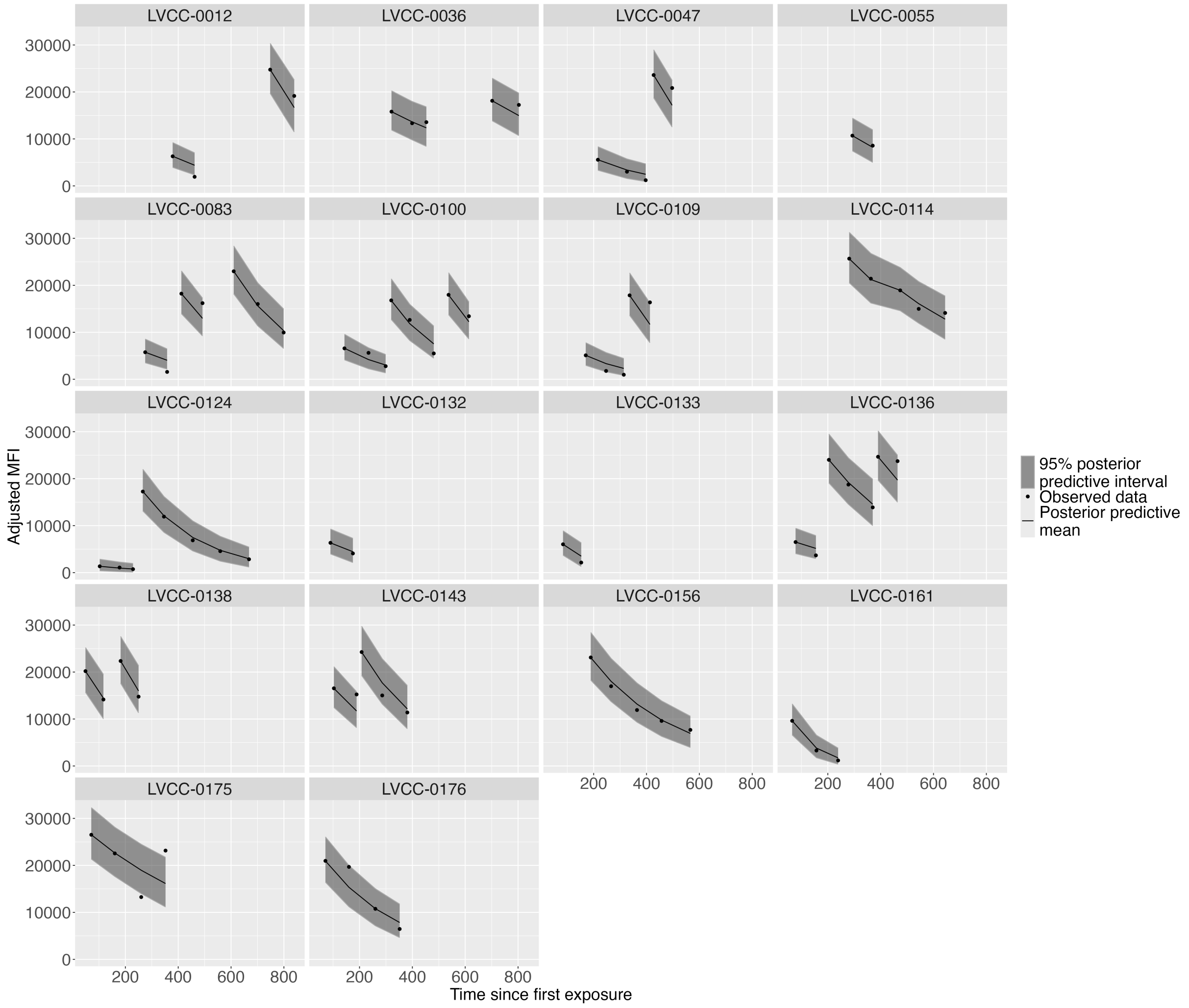


**Figure S20: Single phase decay model fits with observed data for the IgG response to RBD for naive non-mRNA recipients.** Posterior predictive means are represented by the dashed black line and the 95% posterior predictive interval is shown by the grey ribbon. Observed data is indicated by black dots.

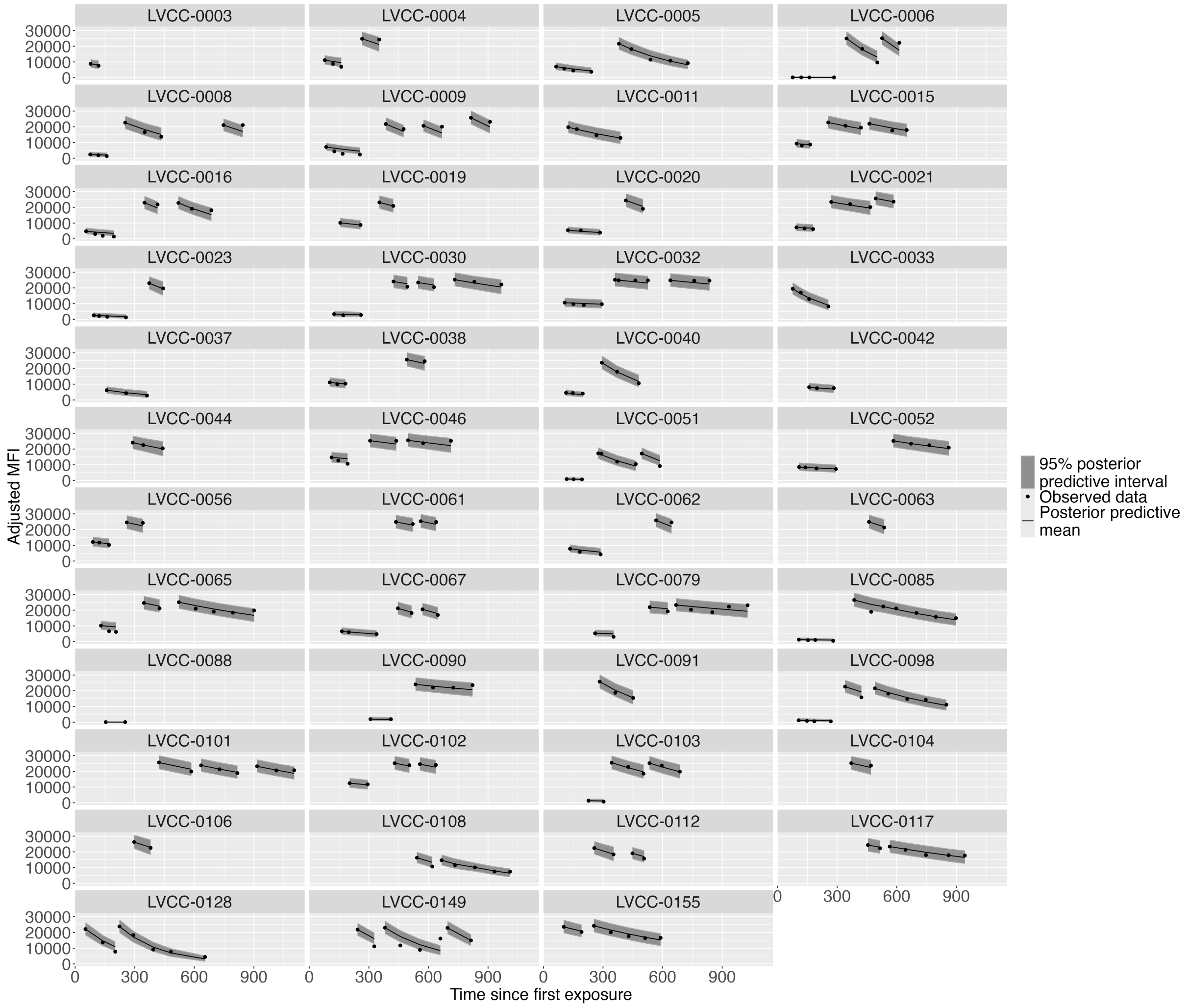


**Figure S21: Single phase decay model fits with observed data for the IgG response to RBD for recovered mRNA recipients.** Posterior predictive means are represented by the dashed black line and the 95% posterior predictive interval is shown by the grey ribbon. Observed data is indicated by black dots.

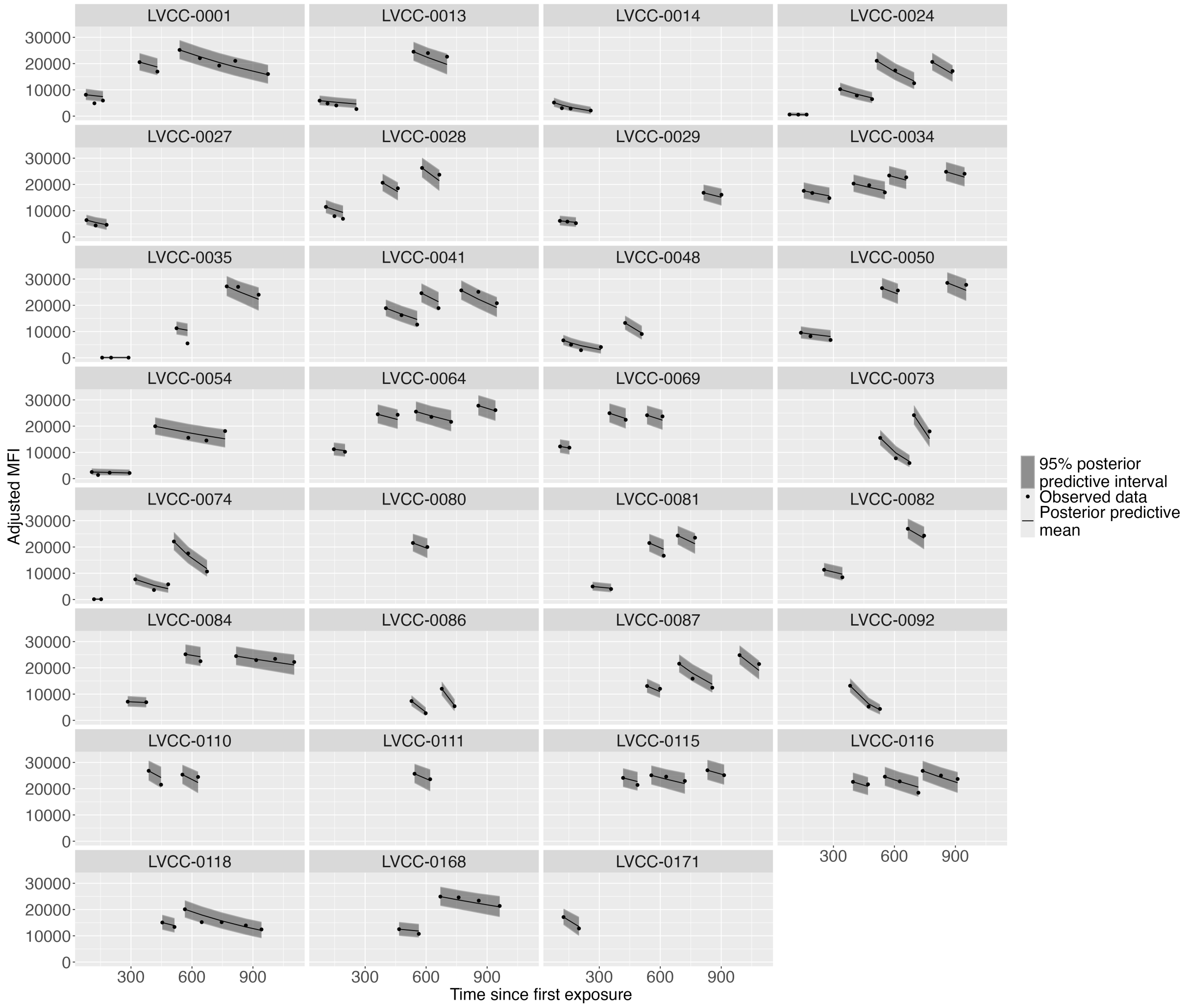


**Figure S22: Single phase decay model fits with observed data for the IgG response to RBD for recovered non-mRNA recipients.** Posterior predictive means are represented by the dashed black line and the 95% posterior predictive interval is shown by the grey ribbon. Observed data is indicated by black dots.

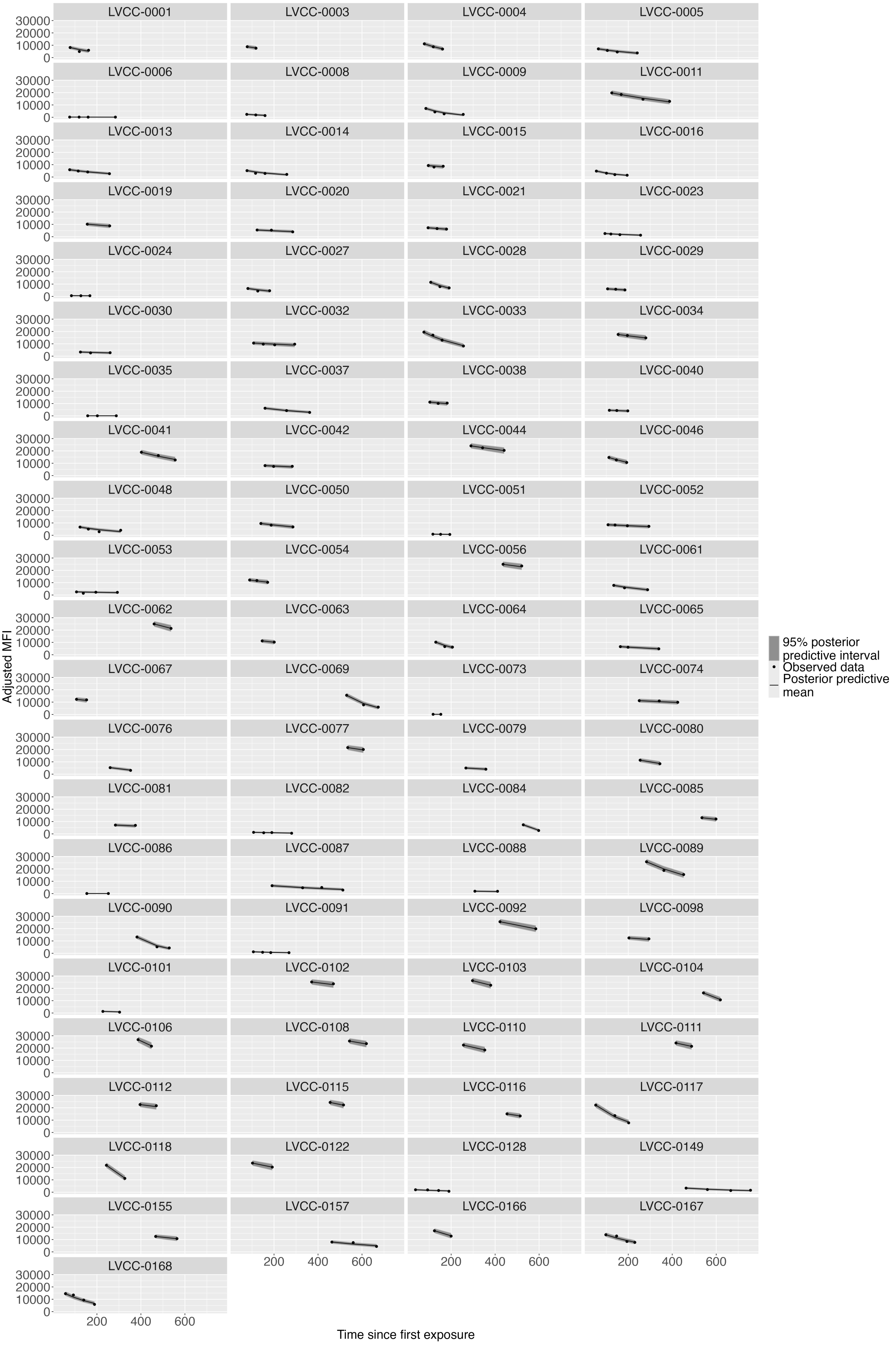


**Figure S23: Single phase decay model fits with observed data for the IgG response to RBD for recovered individuals before vaccination.** Posterior predictive means are represented by the dashed black line and the 95% posterior predictive interval is shown by the grey ribbon. Observed data is indicated by black dots.
